# Systematic Review and Meta-Analysis: Do Youth-Reported Psychosis Symptoms Predict Later Mental Health Diagnosis?

**DOI:** 10.64898/2026.07.13.26357957

**Authors:** Jiya N. Shah, Stephanie H. Ameis, Clarize A. Donato, Isabel Wei, Anisha Miah, Yar Al Dabagh, Kristin Cleverley, Darren B. Courtney, George Foussias, Nicole Kozloff, Aristotle N. Voineskos, Wei Wang, Erin W. Dickie

**Affiliations:** Campbell Family Mental Health Research Institute, Centre for Addiction and Mental Health; Institute of Medical Science, Temerty Faculty of Medicine, University of Toronto; Krembil Centre for Neuroinformatics, Centre for Addiction and Mental Health; Department of Psychiatry, Temerty Faculty of Medicine, University of Toronto; Cundill Centre for Child and Youth Depression, Centre for Addiction and Mental Health; Michael G. DeGroote School of Medicine, McMaster University, Hamilton, Ontario, Canada; Lawrence Bloomberg Faculty of Nursing, University of Toronto; Department of Psychological Clinical Science, University of Toronto Scarborough; Department of Biostatistics and Data Science, University of South Florida, Tampa, Florida, United States

**Author notes:** orresponding author. authors contributed equally (Stephanie H. Ameis, Erin W. Dickie).

## Abstract

**Objective:** Psychosis spectrum symptoms (PSS) are common among children and youth. These symptoms may be clinically significant as studies indicate a heightened risk of mental health disorders, in general, as well as psychotic disorders, specifically, in youth that endorse PSS. This systematic review and meta-analysis investigates the longitudinal association between PSS in children and youth and subsequent mental health diagnosis.

**Methods:** A comprehensive search of Ovid Medline, PsycINFO, and EMBASE databases was conducted to identify longitudinal studies that: (i) assess PSS at a baseline timepoint, (ii) in individuals under 25 years, and (iii) assess mental health disorder diagnosis using a structured assessment at a later time point in the same sample. We conducted a meta-analysis and calculated pooled odds ratios (ORs) for mental health and psychotic disorders using random-effects models. Post-hoc meta-regressions were performed to examine the influence of a number of moderators on the relationship between earlier recorded PSS and subsequent mental health disorders or psychotic disorders.

**Results:** The search yielded 41 eligible studies of which 25 were included in the meta-analysis. Most included studies assessed PSS using brief self-report measures and recruited their samples from clinical or community settings. Among children and youth without an identified mental health diagnosis at baseline assessment, baseline PSS were associated with a 2-fold (OR = 2.07, CI = 1.61 - 2.66, I^2^ = 86.92%, p < 0.0001) increased risk of meeting diagnostic criteria for subsequent mental health disorder diagnosis and a 3-fold increased risk (OR = 3.11, CI = 2.11 - 4.58, (I^2^ = 60.93%, p < 0.0090) of meeting diagnostic criteria for a subsequent psychotic disorder diagnosis with a minimum 1 year follow-up time from baseline assessment. Meta-regression analysis indicated that study quality and sample size explained a substantial proportion of between-study heterogeneity for psychotic disorder outcomes.

**Conclusions:** Our results suggest that administration of simple self-report measures of PSS in both clinical and community settings may be helpful to identify children and youth at higher risk of subsequently meeting criteria for a mental disorder generally, and for a severe mental illness (i.e., psychotic disorder), specifically. Future longitudinal studies should focus on improving study design characteristics to increase confidence in identified longitudinal associations. The results of our work suggests that integration of self-report measures of PSS may be useful in a variety of settings to identify youth at increased risk of subsequent mental illness.

## Introduction

Mental health disorders represent a substantial burden of disease, contributing significantly to morbidity, disability, and reduced quality of life^1^. A large proportion of mental health disorders emerge early in life, with onset occurring in 35% of individuals before age 14 and 63% before age 25^2^. The perceived need for youth mental health support has risen in recent decades^3^, such that available health care systems and resources cannot keep up with demand, resulting in long wait times for specialty mental health care^4^.

Given the prevalence and potential long-term impact of youth-onset mental health disorders, efforts to identify easily measurable markers of elevated long term mental health risk are critical to directing limited youth mental health care resources to those at greatest need. Risk states for psychosis have been well characterized through constructs such as clinical high-risk (CHR), ultra-high risk (UHR), and at-risk mental states (ARMS), which aim to identify individuals in prodromal stages of psychosis^5^. Meta-analyses conducted on this population have reported that 25% of individuals meeting ‘at-risk’ criteria transition to developing a diagnosable psychotic disorder within 2-3 years and are more likely to meet criteria for co-occurring mental health disorders^6^. While identification of individuals meeting CHR criteria is important for resource allocation^7,8^, CHR identification requires individuals to present to clinical settings for resource intensive assessments to determine CHR status^9^. Thus, additional approaches are still required to identify those youth at greatest risk for poor mental health outcomes and in need of specialty care in the general population; ideally, such approaches would add minimal burden/expense to the busy care systems that aim to support them.

In this context, easy to administer self-report assessments for measures such as subclinical psychotic symptoms or psychosis spectrum symptoms (PSS) may fill an important gap. The term PSS encompasses psychotic experiences (PEs) and psychotic like experiences (PLEs) and describes attenuated, schizophrenia-like symptoms such as unusual thoughts, perceptions, and beliefs that do not meet full criteria for a psychotic disorder. Such experiences have been found to occur in 17% of children and 7.5% of adolescents in the general population^10^. A retrospective study indicated that brief or attenuated psychotic symptoms (in the absence of a primary persistent psychotic disorder) can be present years before a young person meets criteria for a psychotic disorder, suggesting that PSS may be an important risk marker^11^. The presence of PSS has been associated with reduced neurocognitive and global functioning, increased likelihood of psychiatric diagnosis, substance use, and suicidal ideation^12^. If PSS assessments can identify children and youth at risk for poor mental health outcomes at a large scale in the community, they may be able to simplify and expedite identification and triage of youth with higher mental health care needs. This is particularly true if PSS identification can add predictive value for poor mental health outcomes (e.g., over and above other mental health symptoms or functional measures).

Two meta-analyses to date have synthesized the literature examining the association between reported PSS in children and youth and risk of subsequent mental health and psychotic disorders. Healy and colleagues (2019) reviewed 5 cross-sectional and longitudinal cohort studies examining community-based samples of children and youth (<18 years old). They conducted a pooled meta-analysis of longitudinal studies (k =7, n ∼1600) and found that the presence of childhood psychotic experiences was associated with a three-fold increased risk of a young person meeting criteria for any subsequent mental disorder and four-fold risk of primary persistent psychotic disorder diagnosis (k = 5)^13^. Burton and colleagues (2024) reported in their meta-analysis of prospective community cohort studies (k=5, n∼16,000) that individuals with reported PSS had a four-fold increased longitudinal risk of primary persistent psychotic disorders^14^.

While informative, previous meta-analyses did not examine whether PSS risk is similar in mental health help-seeking versus community ascertained populations, and limited sampling to those <18 years of age. Thus, excluding older youth (i.e., >18 years) who are at higher risk of developing a psychotic disorder as they are approaching peak age of psychotic disorder onset^15^. It is critical to better understand whether PSS in help seeking populations indicate elevated risk for poor outcomes as approximately two thirds of those presenting with a psychotic episode between the ages of 15 and 29 have presented for mental health support in the 3 years before formal diagnosis^16^. In addition, to our knowledge, no study has examined whether factors associated with long-term mental health risk (e.g., follow-up after PSS assessment, cannabis use, type of PSS assessment, and sex distribution) influences the association between PSS and diagnostic outcomes.

This review extends previous meta-analyses exploring the association between self-reported PSS and risk for subsequent mental health diagnosis by including primary studies with help seeking samples in addition to community samples, youth up to age 25, and integrating longitudinal studies published since the most recent meta-analysis. The primary objective of this review is to investigate the longitudinal association between baseline PSS during childhood and youth and emergent or persistent mental health disorder diagnosis at prospective follow-up. The secondary aim is to examine whether PSS are differentially associated with an increased risk of specific categories of mental health disorders, with an emphasis on risk for psychotic disorders.

## Methods

### Search Strategies

A systematic review was conducted of all published literature on PSS in children and youths and subsequent mental health disorder diagnosis following the Preferred Reporting Items for Systematic Review and Meta-Analysis (PRISMA) reporting guidelines for systematic reviews (Page et al. 2020). The search was conducted in Ovid Medline, PsycINFO, Web of Science, and EMBASE electronic databases, from their inception to March 2025. The following search strategy was developed with the assistance of a librarian: 1. ((psychosis or psychoses or psychotic) adj3 (symptom* or experience*)).ti,ab. 2. cohort analysis’/ or ‘longitudinal study’/ or ‘follow up’/ **OR** ((cohort or concurrent or incidence or ‘follow up’ or longitudinal) adj2 (stud* or analys* or survey*)).ti,ab. 3. (teen* or youth* or adolescen* or juvenile* or child* or (young adj2 (adult* or person* or individual* or people* or population* or man or men or women)) or youngster* or first-grader* or second-grader* or third-grader* or fourth-grader* or fifth-grader* or sixth-grader* or seventh-grader* or highschool* or college* or ((secondary or high*) adj2 (school* or education))).ti,ab. or ‘adolescent’/ or ‘young adult’/(Lacourse n.d.). Any duplicates were handled by Covidence which is a web-based platform that streamlines the production and collaboration on systematic reviews^17^.

### Eligibility Criteria

Studies were eligible if they were published in peer-reviewed journals, written in English, and assessed PSS at baseline in participants with the sample average age of < 25 years old. The studies had a minimum follow-up time of at least one year duration between the timing of initial PSS measurement to subsequent assessment of diagnostic outcomes and the longest time point was used in the event that they had multiple time points. In this review, PSS is defined as experiencing any of the following: brief psychotic experiences (delusions, hallucinations, thought disorganization), persistent psychotic experiences, psychotic like experiences (attenuated delusions or perceptual abnormalities), attenuated disorganization or attenuated negative symptoms^12,18,19^. PSS could have been identified through self-report questionnaires, structured research interviews, or clinical interviews. Studies that included higher risk for psychosis samples such as those with family history of psychotic disorder, meeting CHR criteria, and seeking treatment for other mental health disorders^20^ were included. Both studies of community samples or individuals recruited through mental health services^20^ were included. For the outcome, studies must ascertain diagnostic outcomes using standardized criteria from any edition of the Diagnostic and Statistical Manual of Mental Disorders (DSM)^21^ or the International Classification of Diseases (ICD)^22^ collected via established semi-structured interviews conducted by trained assessors. We excluded studies of participants with a threshold primary psychotic disorder diagnosis at study inception, studies that only determined prior PSS or diagnosis at follow-up retrospectively (including cross-sectional studies), controlled clinical trials, case series, dissertations, abstracts, grey literature, and study protocols.

### Data Collection Process

The Covidence platform was used to conduct the title and abstract and full text screening^17^. Title and abstracts were independently screened in duplicate by two of four reviewers (JS, CD, YA, AM). Full text articles were screened in duplicate by 3 reviewers (JS, CD, IW). Any discrepancies were resolved by a third reviewer (one of JS, CD, IW).

### Quality Assessment

An assessment of study quality was conducted using a modified Newcastle-Ottawa Scale for Cohort Studies^23^. This scale was used to assess the quality of the studies as well as risk of bias across included studies; scores ranged from low to high on a scale of 1-8 (Supplemental Table 1)The quality assessment was conducted independently in duplicate by both JS and IW and any discrepancies were discussed and resolved amongst the two reviewers until a consensus was reached.

### Data Items and Metrics

Data was extracted by JS and IW independently and discrepancies were resolved by consensus. Quantitative data extraction included sample size, mean age at baseline, and follow up time where we extracted the longest follow up time available. For qualitative data, we extracted the sample type (higher risk or community sample), PSS threshold criteria and assessment, the mental health diagnostic criteria and assessment at follow up, and the type of outcomes available.

Unadjusted odds ratios (ORs) were extracted when available from the studies. If the ORs were not found within the study, ORs were calculated using data presented in the study, where possible. In the event that ORs were not present and there was insufficient data to calculate them, adjusted ORs were used. Authors were also contacted if they presented other metrics within their study such as risk ratios (RRs) or hazard ratios (HRs) to request data to calculate ORs.

### Statistical Analysis

All data analysis was conducted in R version 4.6.0 (R Foundation for Statistical Computing)^24^ using the metafor package version 4.8^25^. To account for potential variability across studies, we conducted random-effects models. Heterogeneity across studies was quantified using the I² statistic, with thresholds of 25%, 50%, and 75% representing low, moderate, and high levels of heterogeneity, respectively^26^. For the primary outcome of any future mental health disorder diagnosis, where studies reported a risk estimate for “any mental health diagnosis,” we extracted random effects pooled ORs (OR Model 1), with estimates of heterogeneity across the studies. In the event that a study reported multiple diagnoses, we conducted within-study fixed-effect pooling of individual diagnoses to create a single, overall mental health disorder diagnosis OR. If a study reported a specific type of mental health disorder diagnosis, it was included in the primary outcome of any future mental health disorder. Potential outliers and influential studies were assessed using absolute studentized residuals above 3 and based on leave-one-out and combinatorial meta-analyses^27^ and removed from the meta-analyses. A sensitivity analysis was conducted with the outliers included as well (Supplemental Figure 2 and 4). Publication bias was assessed by examining funnel plots and Egger regression tests for funnel plot asymmetry^28^. Additionally, we reported the population attributable fraction (PAF) for studies with sufficient information on baseline PSS prevalence rates. PAF estimates the proportion of disorders in the population that may be attributable to childhood and adolescent PSS, assuming the observed association is causal^29^. Where possible, we also extracted diagnosis specific risk estimates and conducted meta-analysis to examine whether earlier recorded PSS was associated with specific subcategories of mental health disorders including psychotic disorders (OR Model 2), anxiety/stress disorders, depressive disorders, and substance use disorders.

### Meta-regression Analysis

Post-hoc meta-regressions were performed where the number of studies available were sufficient (k ≥ 10), to examine the influence of a number of moderators on the relationship between earlier recorded PSS and subsequent mental health disorders or psychotic disorders. Independent variables that we wished to examine based on the availability of adequate studies included: average age at baseline assessment, follow up time period, sample size of a given study, cannabis use reported in the sample, sex distribution, and PSS assessment type (self-reported questionnaires versus clinician administered or rater administered interviews).

## Results

### Study Selection

The search yielded 8511 studies, and after removing duplicates, 5286 titles and abstracts were reviewed for relevance. This resulted in 237 studies being identified for full text screening. Based on the inclusion and exclusion criteria, 41/237 met criteria for inclusion in the review. Of those included, 25 studies were included in the meta-analysis as they had sufficient data for analysis (Table 1). Three authors were contacted to request the additional measures needed to calculate the ORs^55,68,70^. Reasons for exclusion after full text screening are provided in the PRISMA flow diagram^71^(Figure 1).

**Figure 1.**
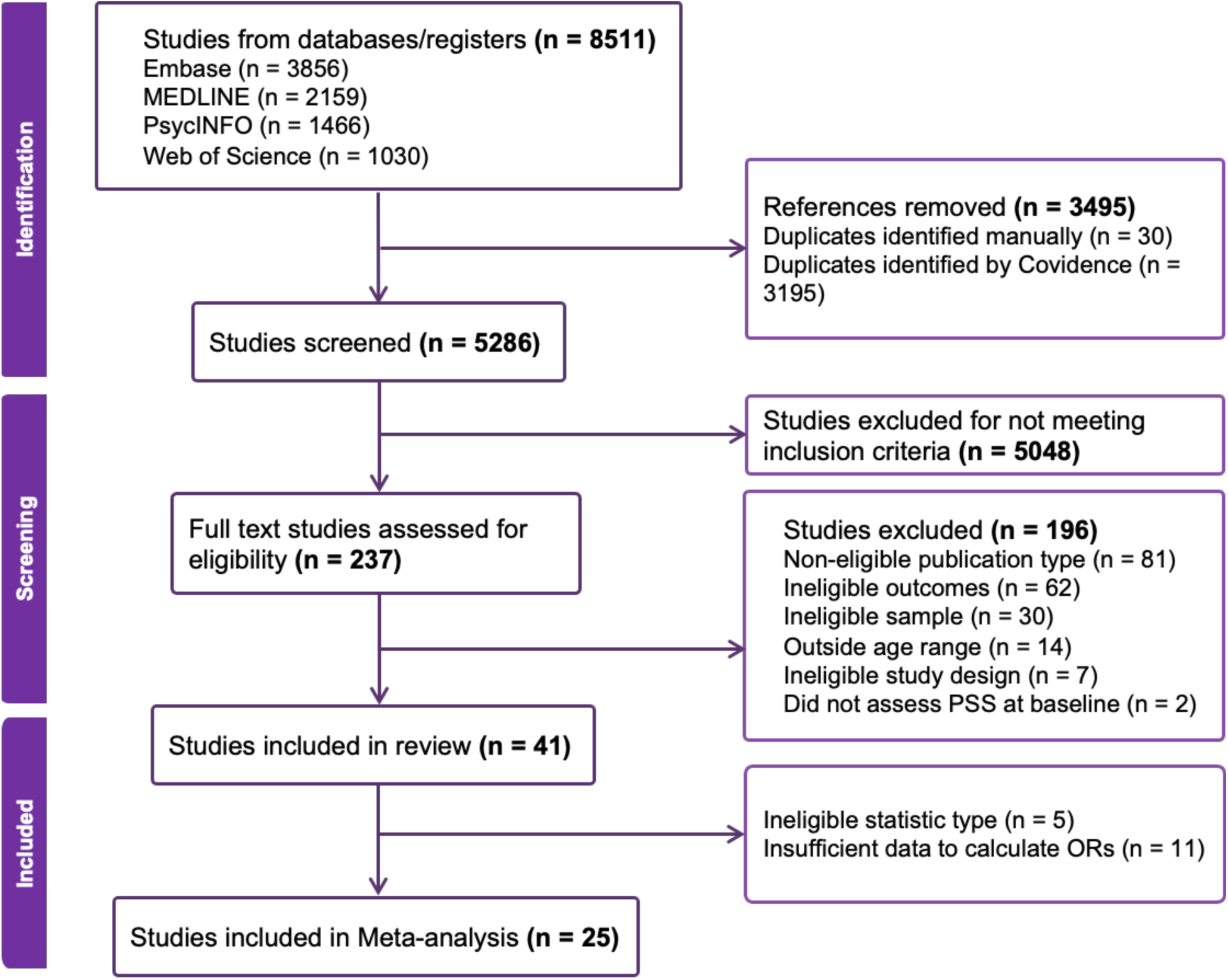
PRISMA flow diagram for study inclusion.

**Figure 2.**
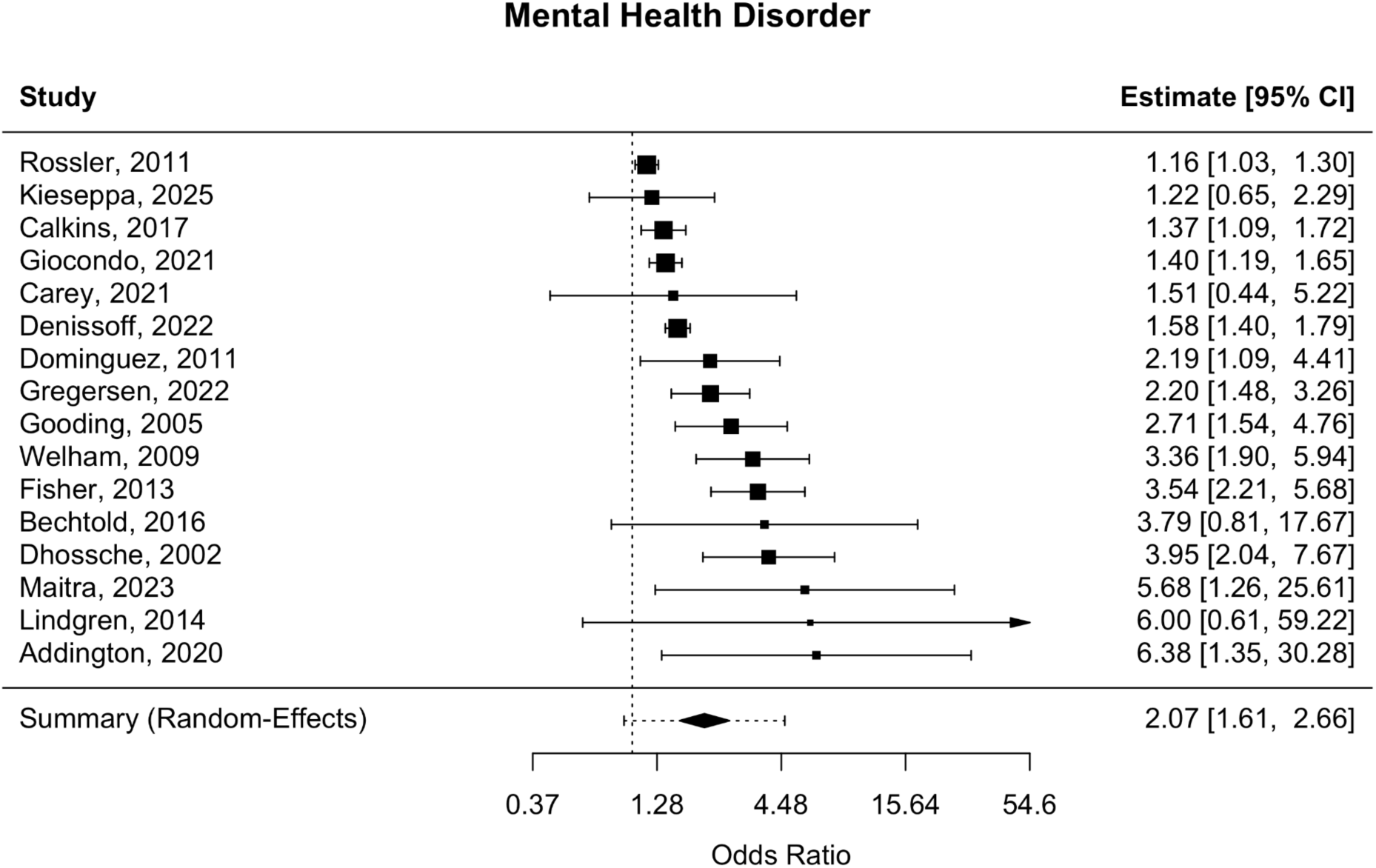
Forest Plot of Odds Ratios for Mental Health Disorders as the Outcome.

**Figure 3.**
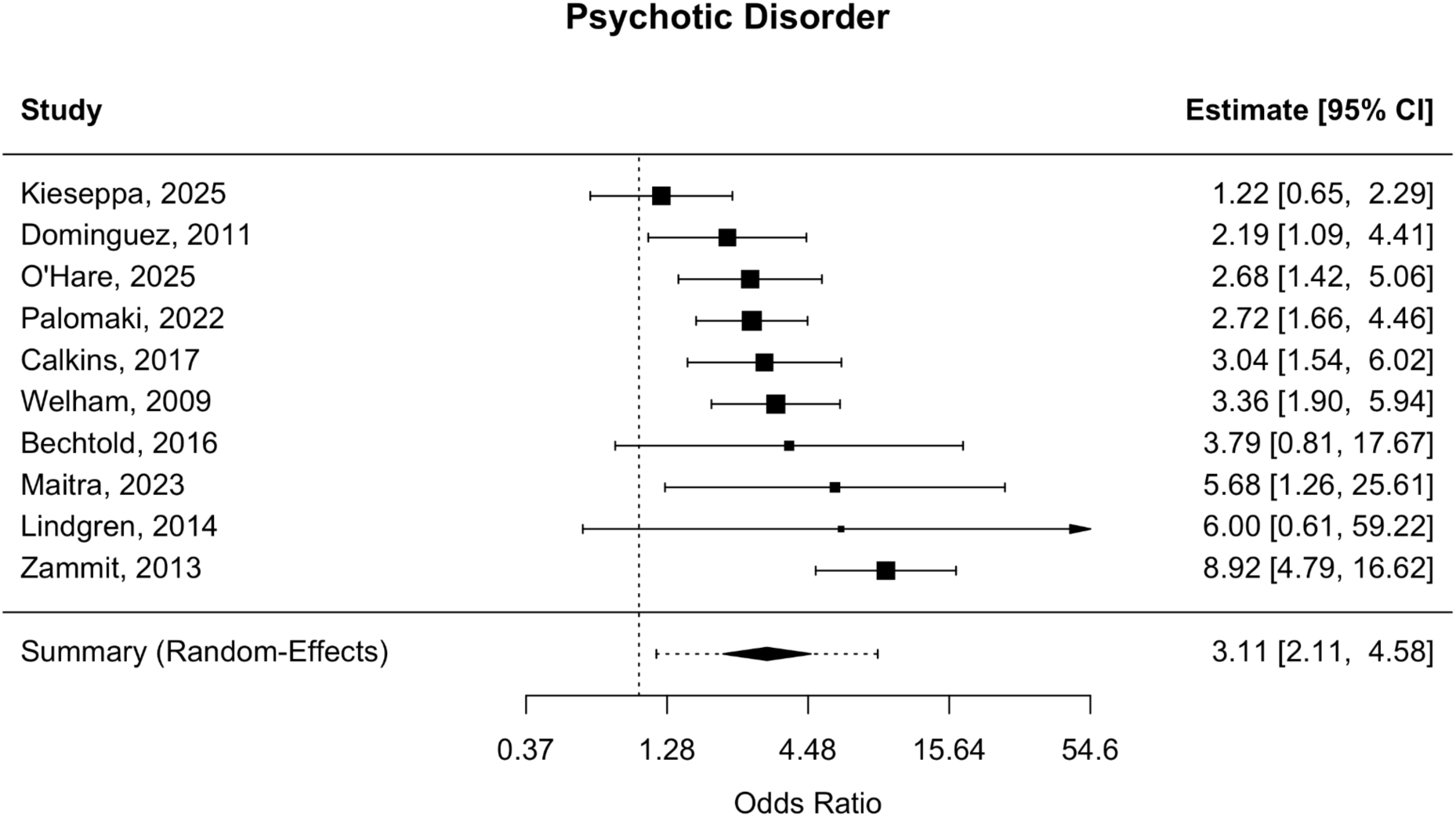
Forest Plot of Odds Ratios for Psychotic Disorders as the Outcome.

**Table 1.**
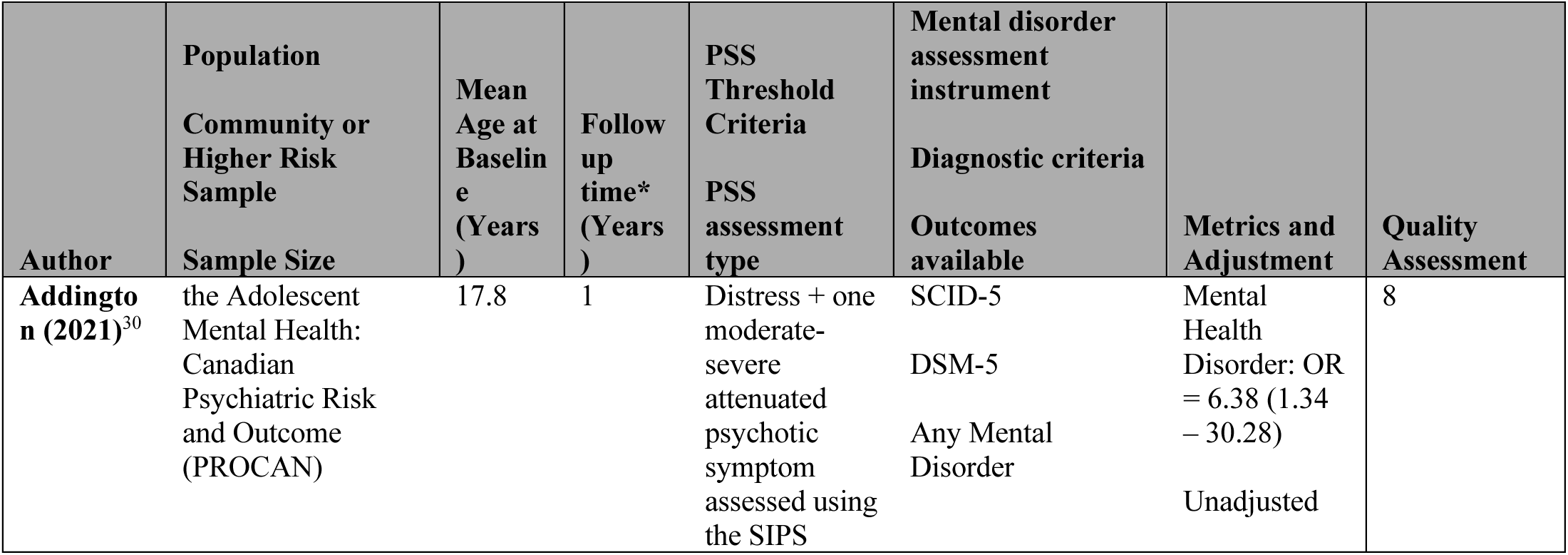
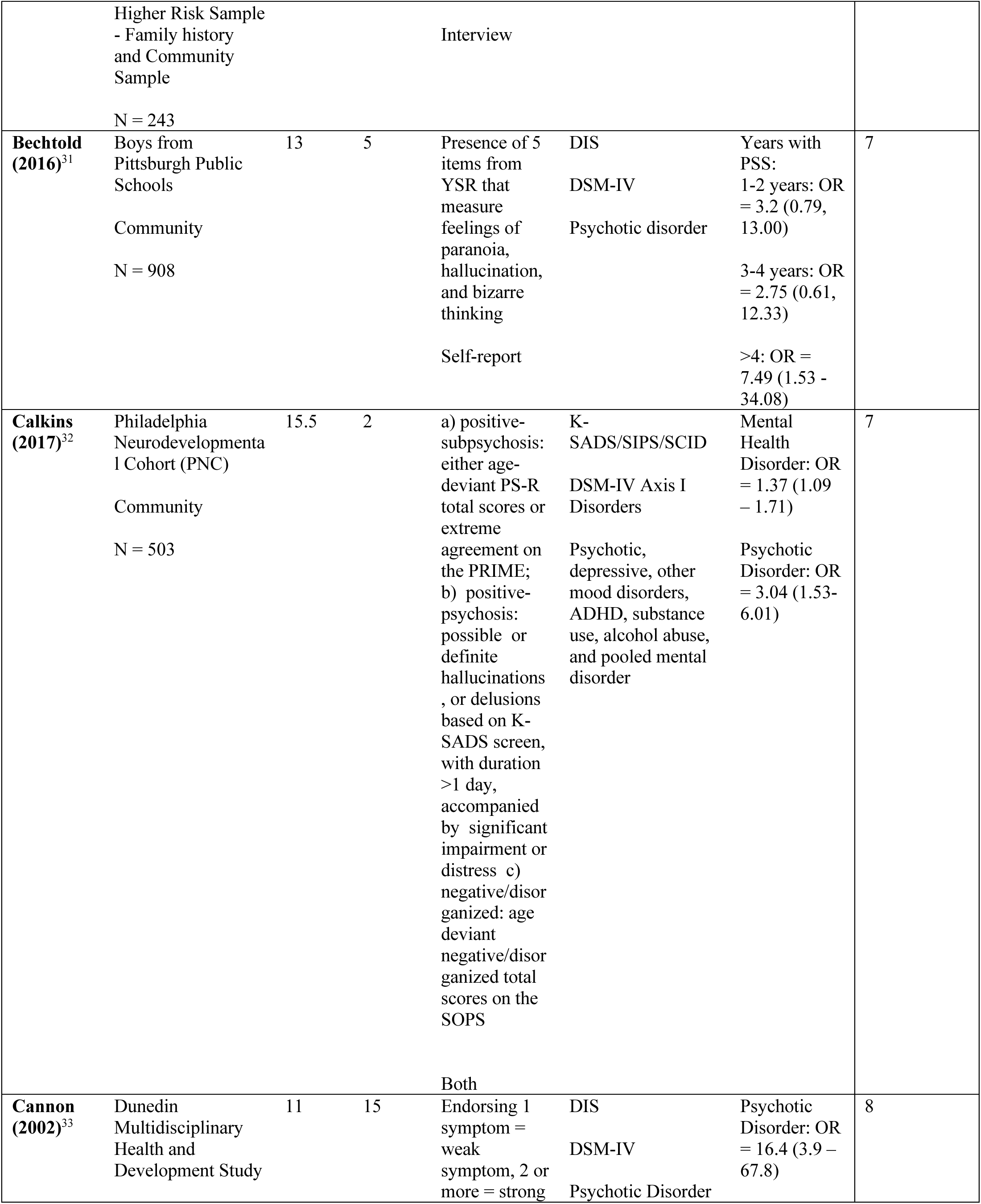
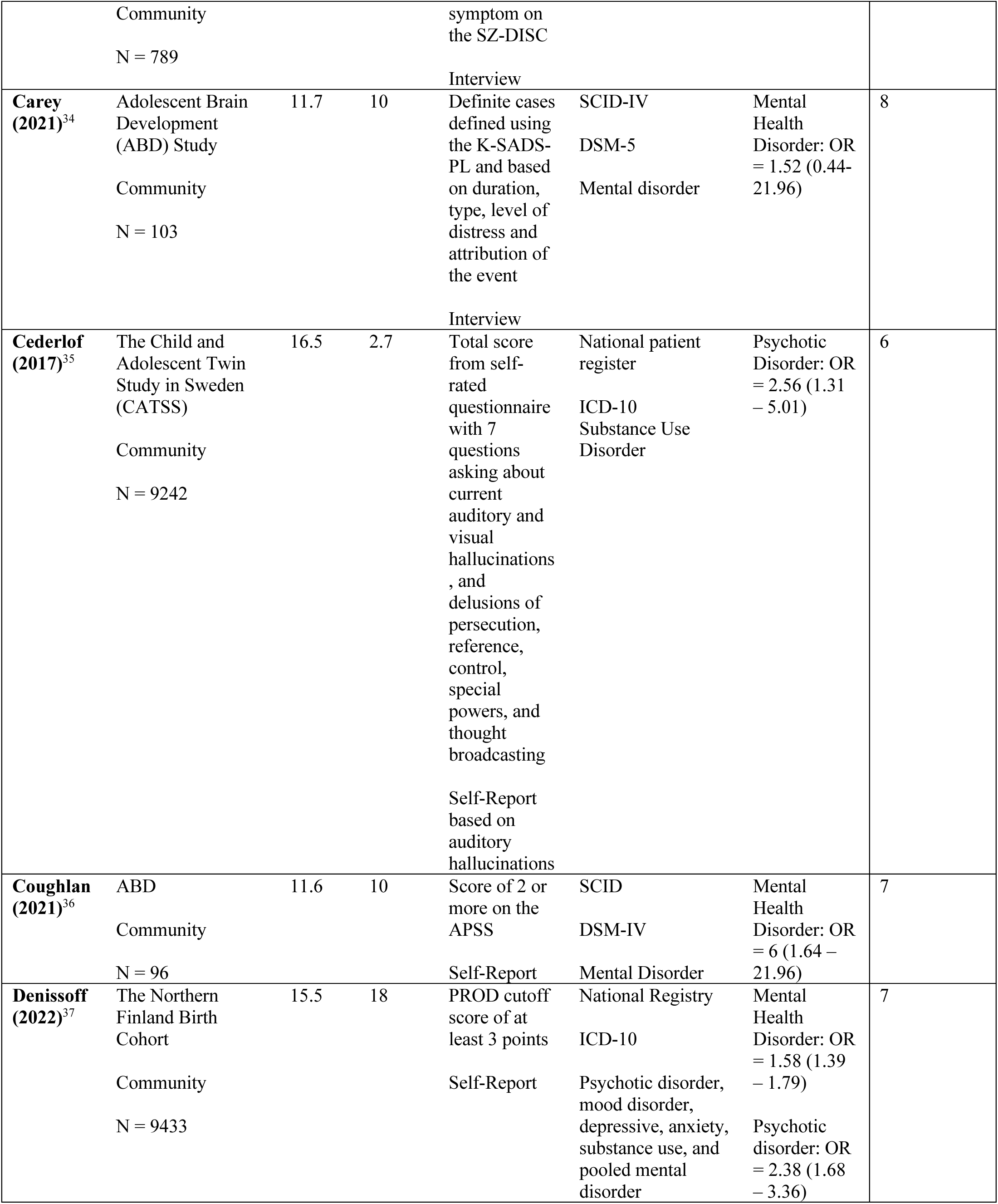
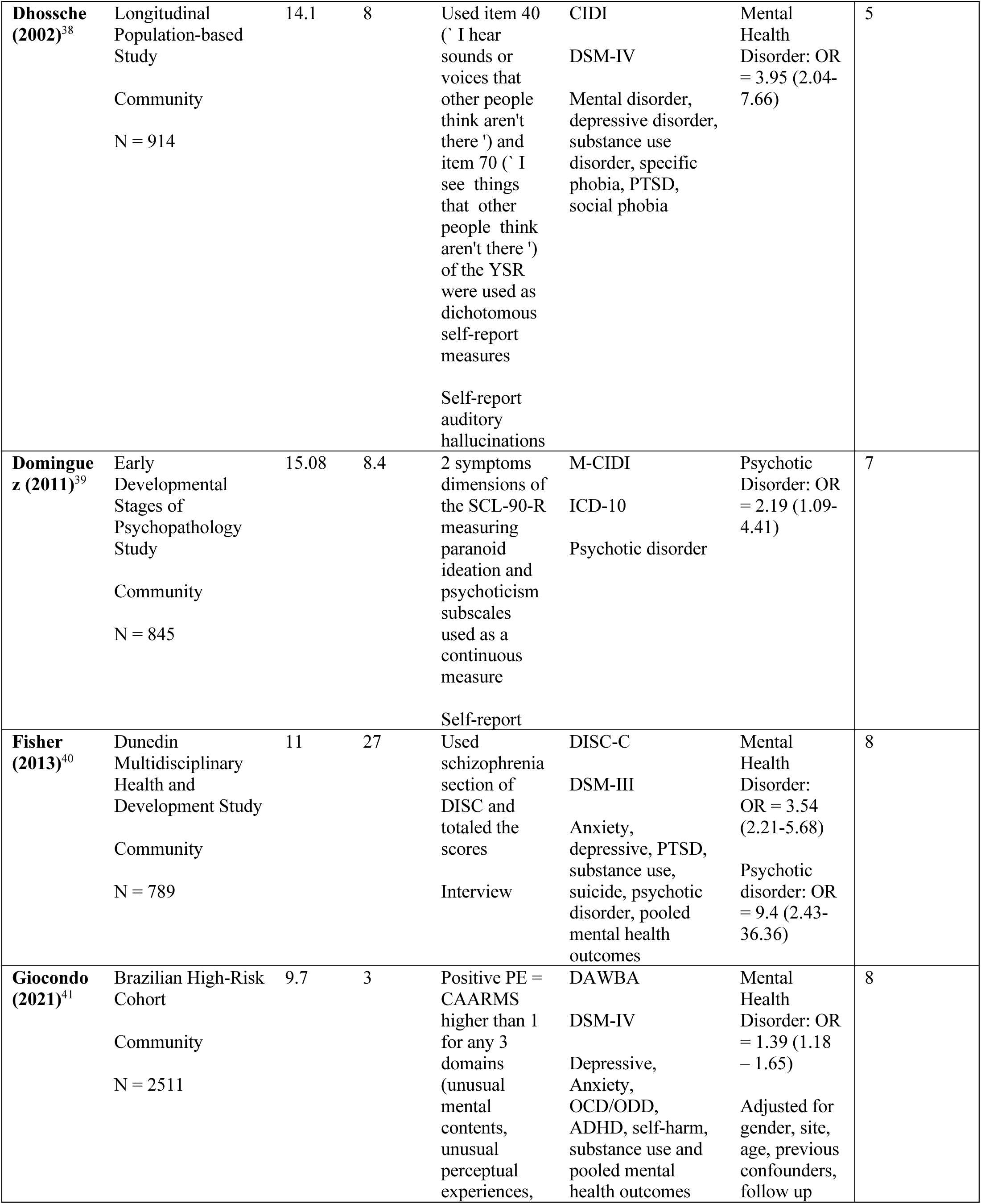
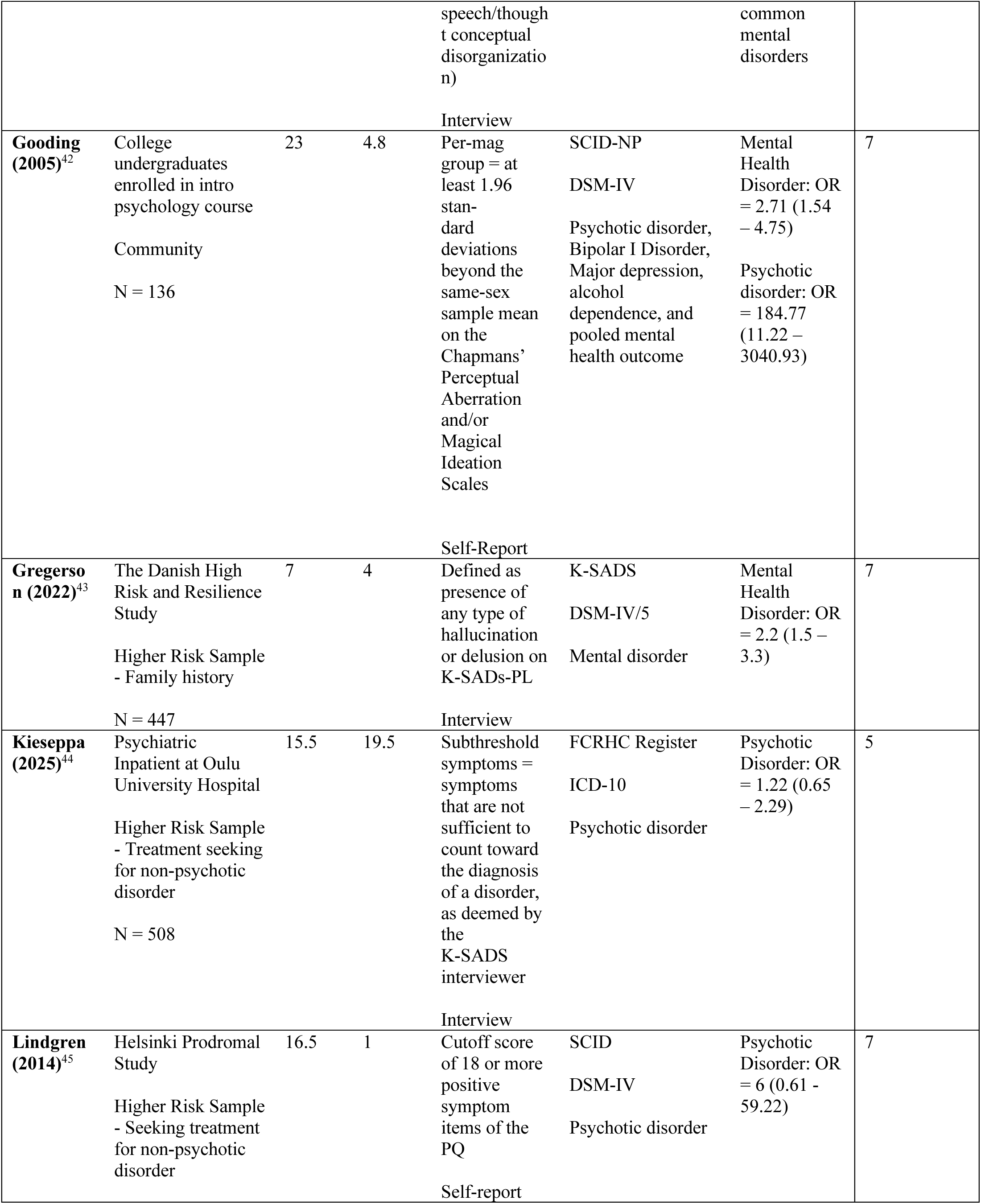
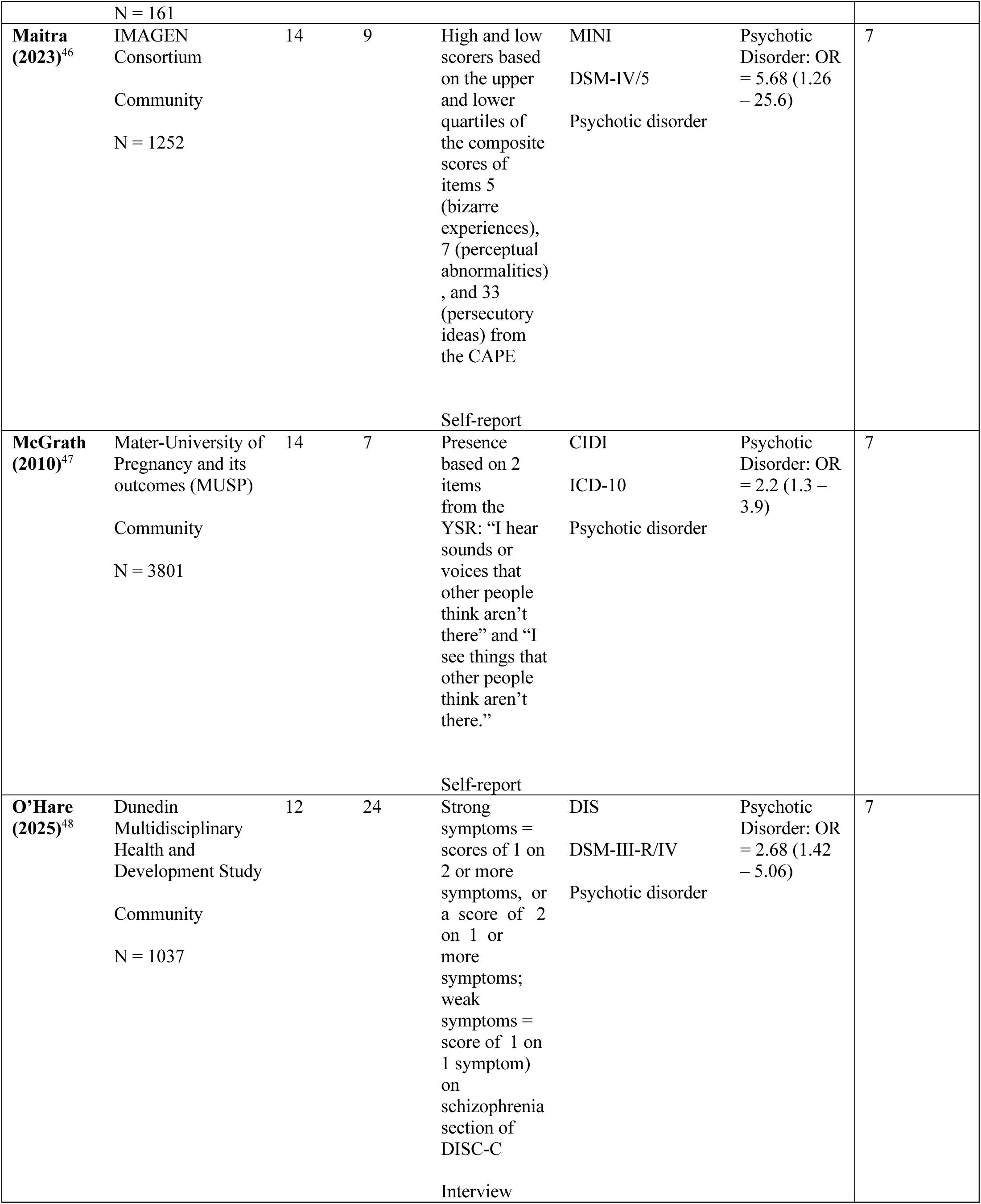
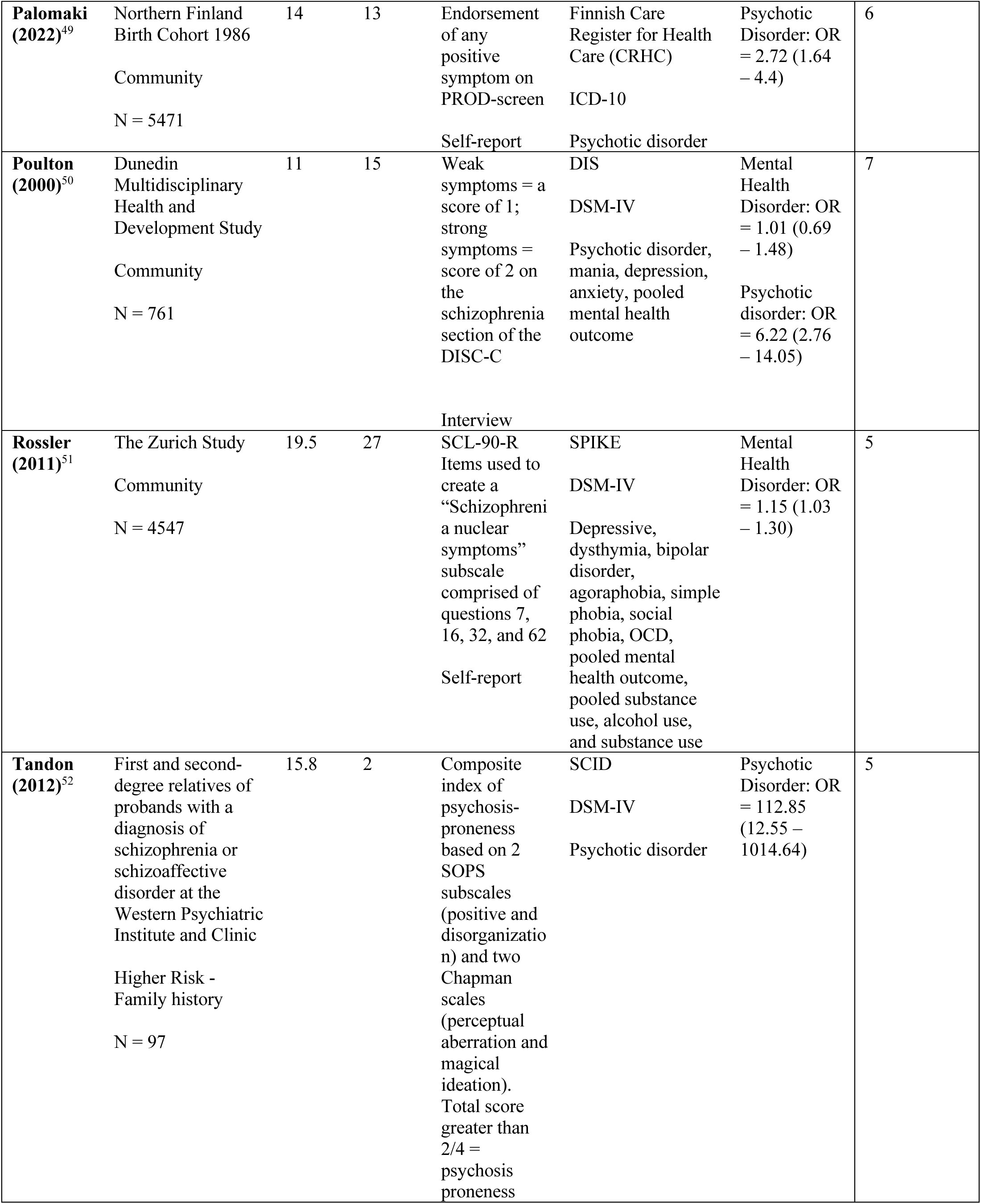
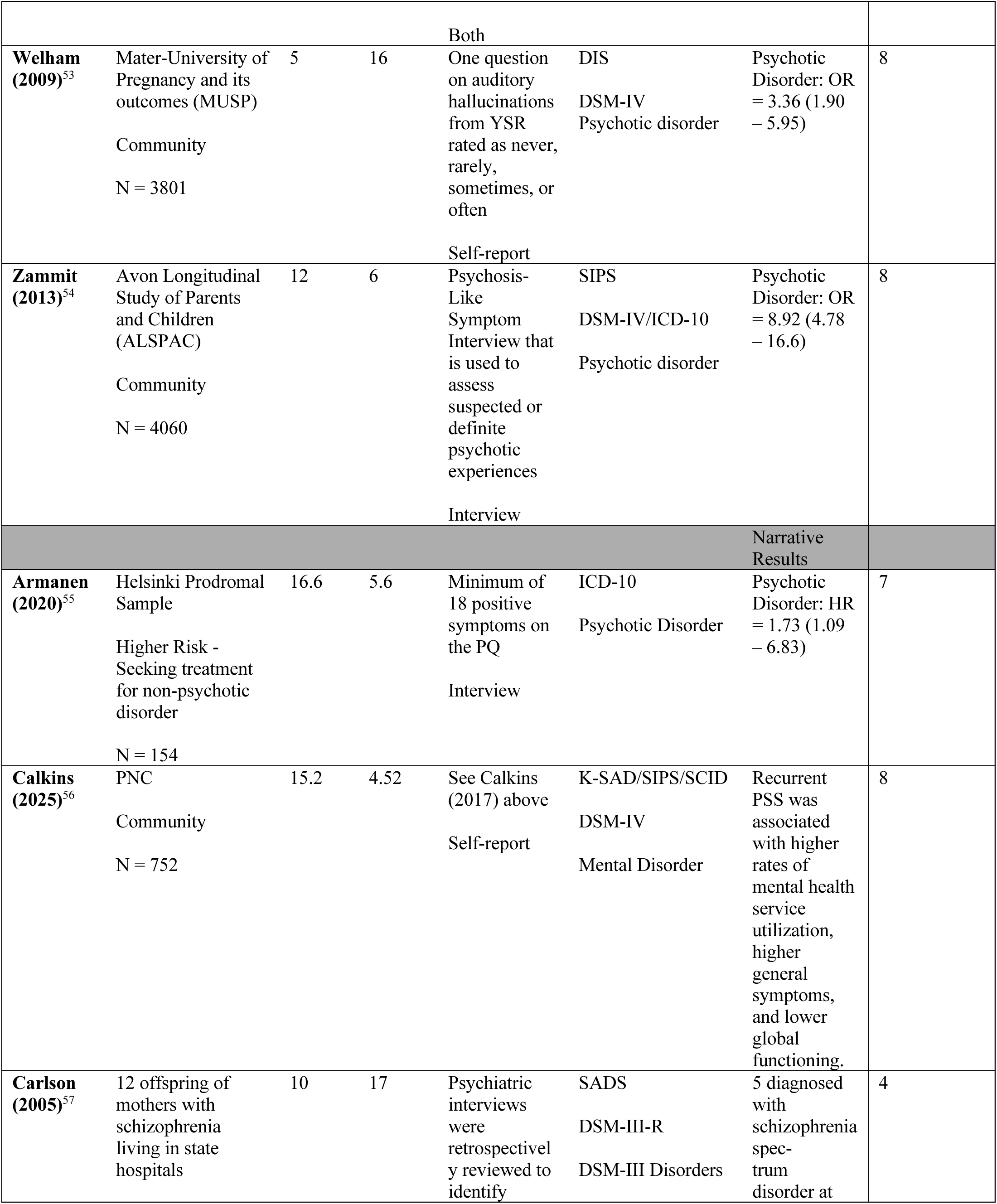
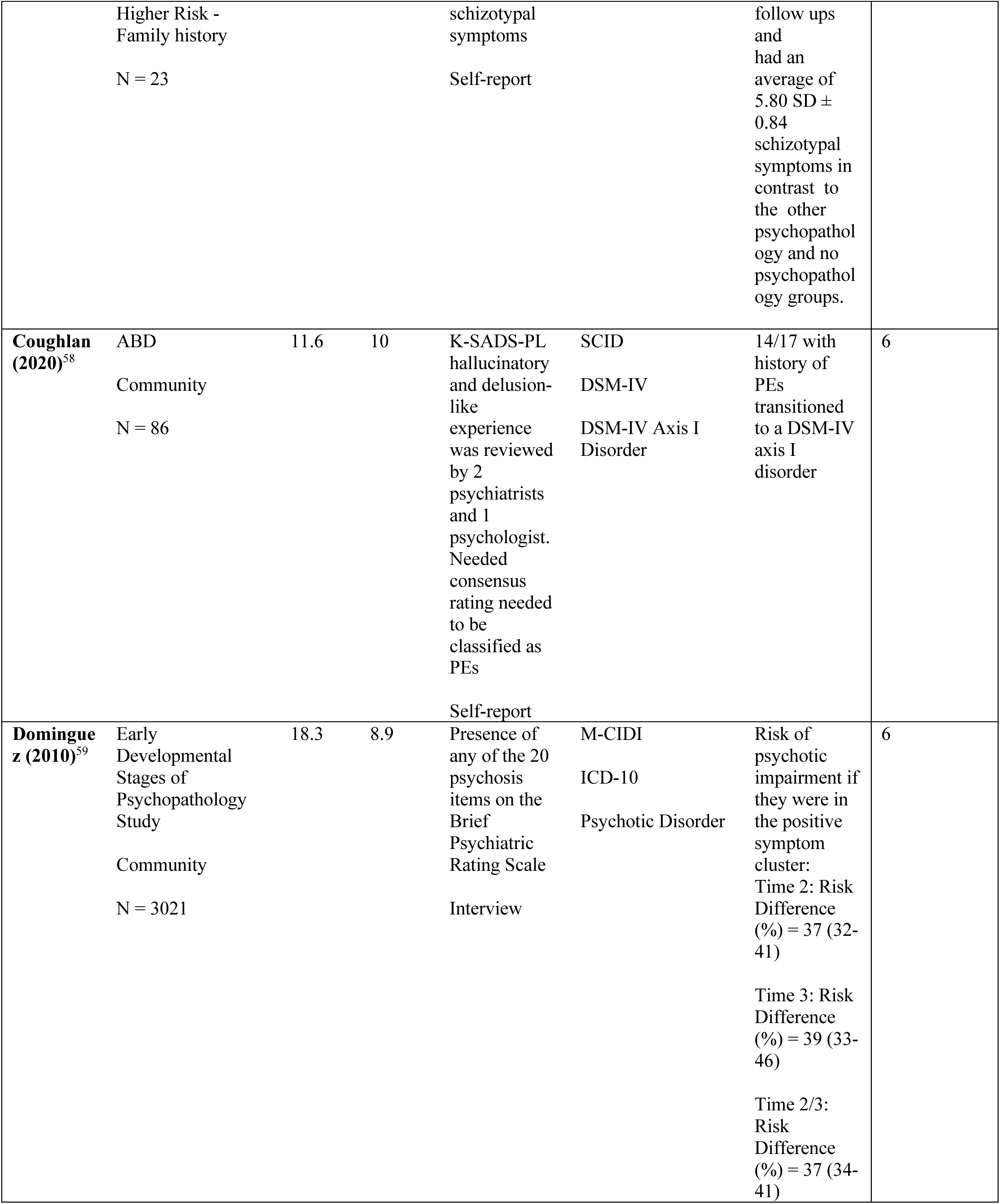
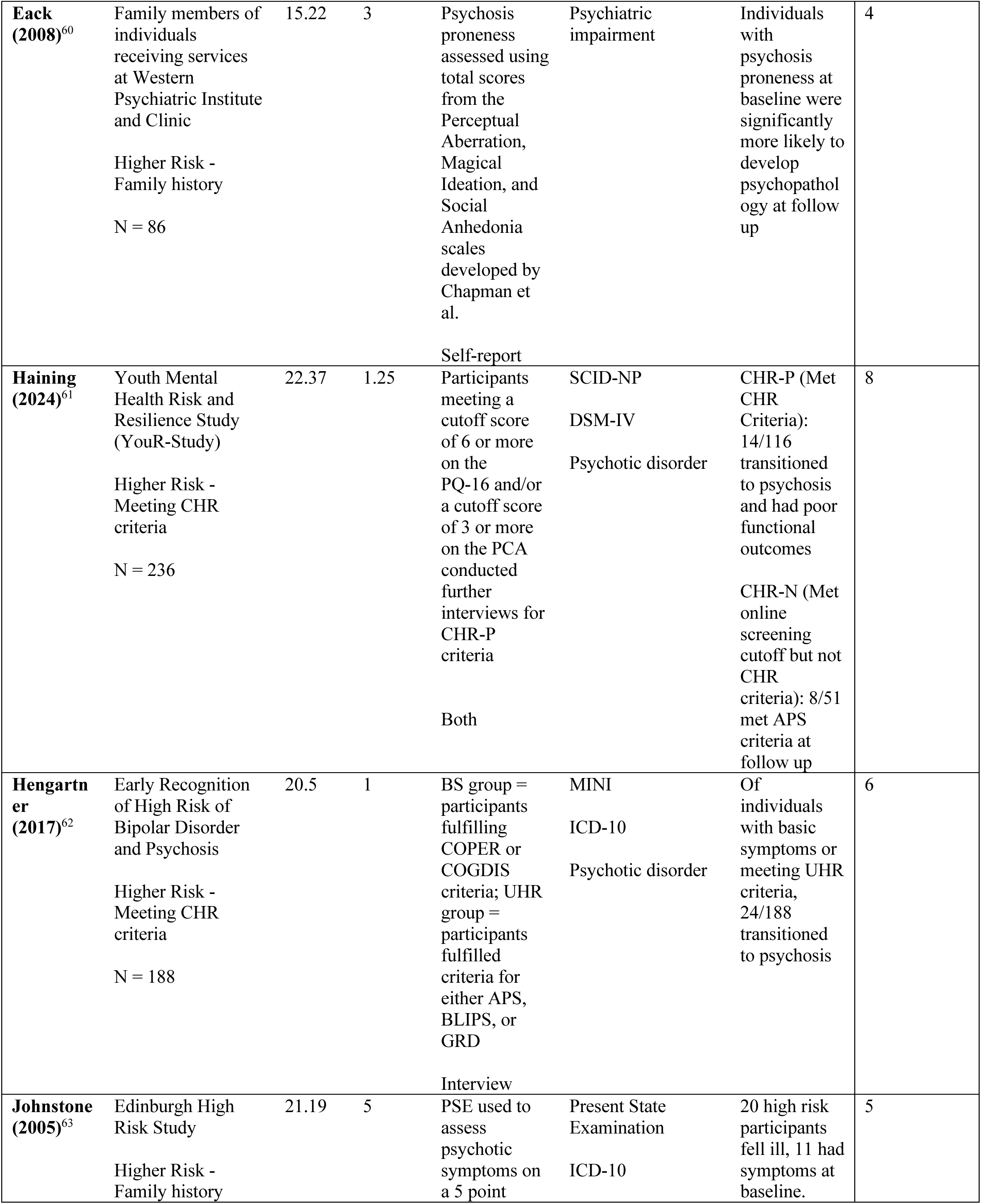
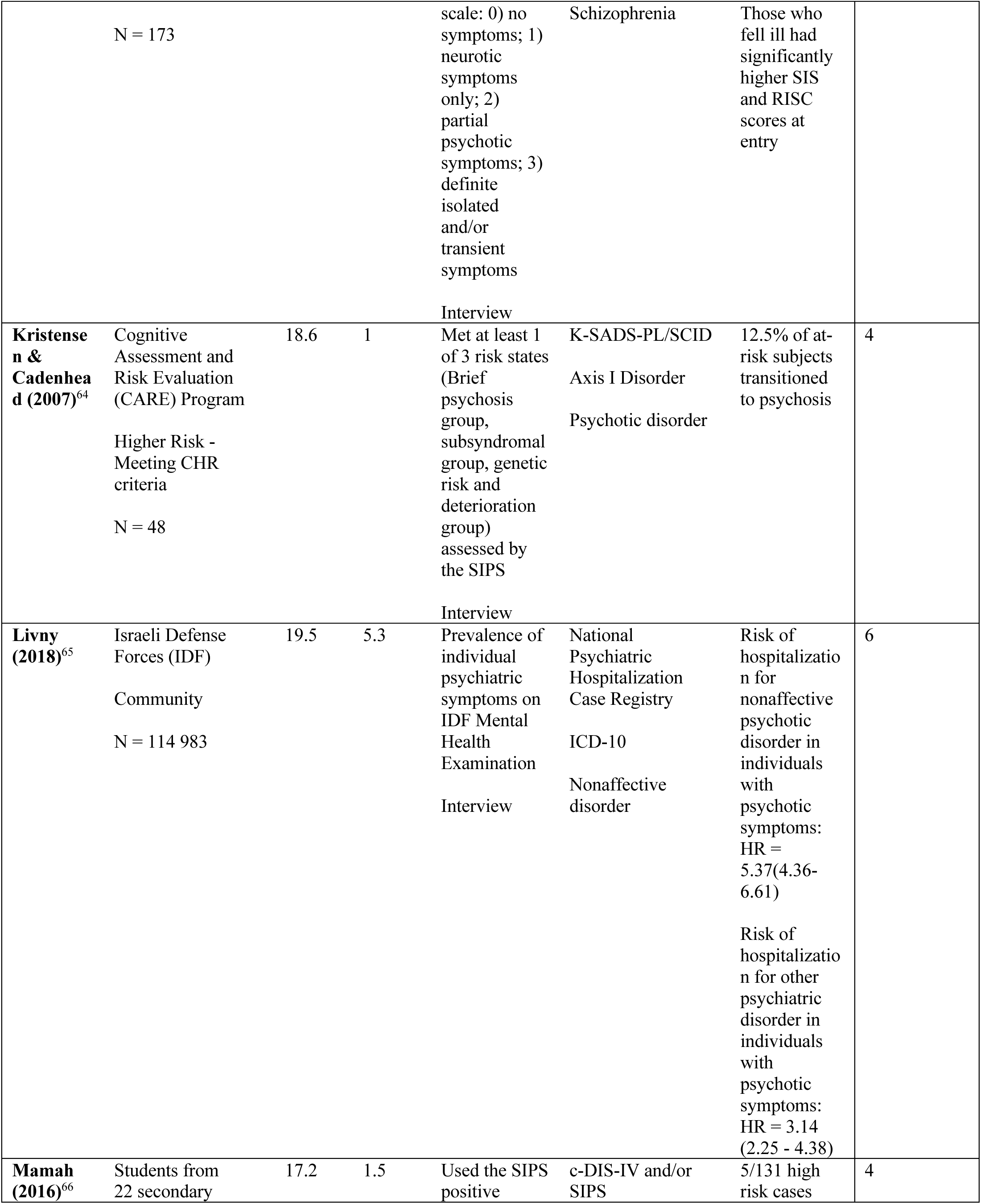
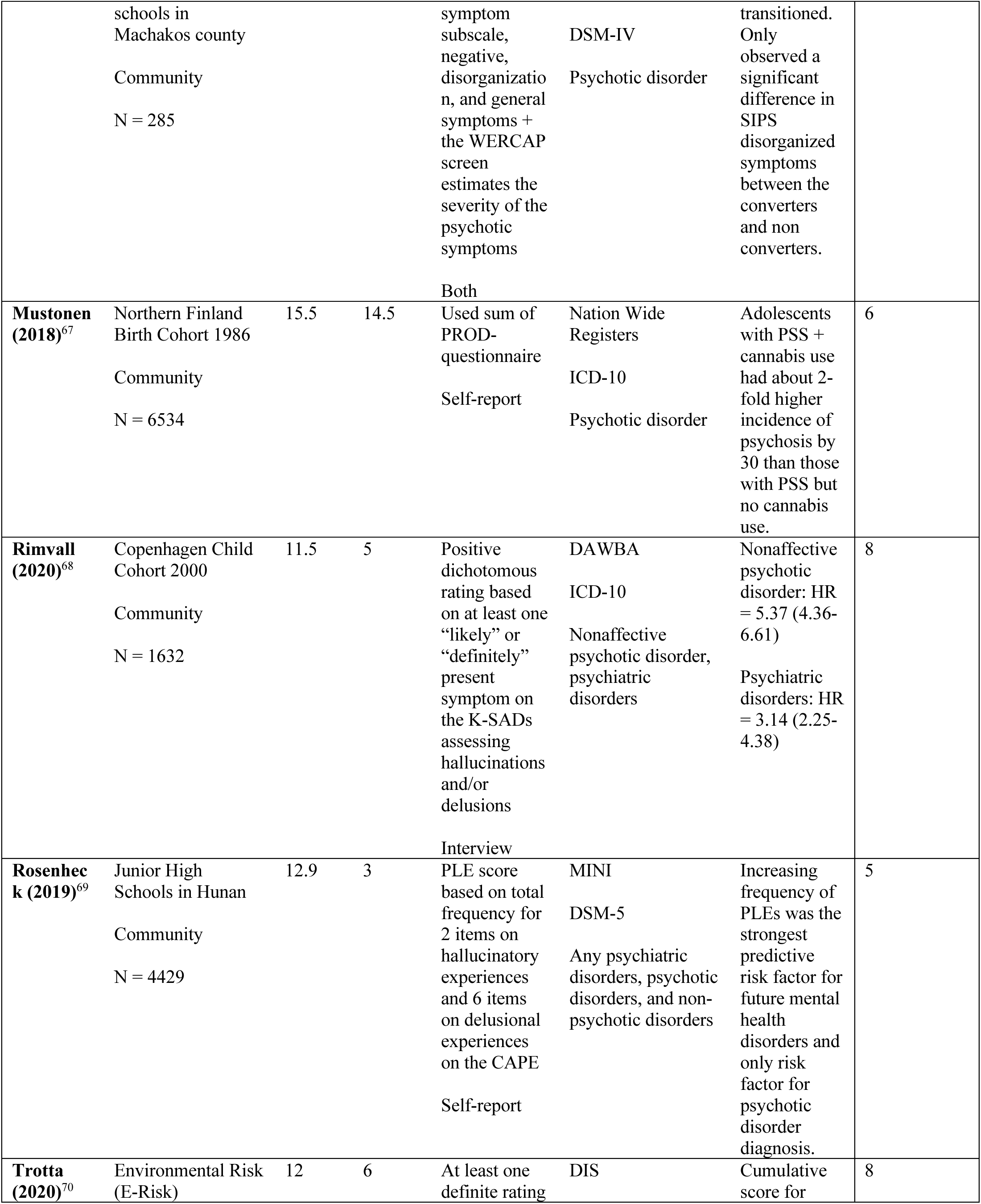
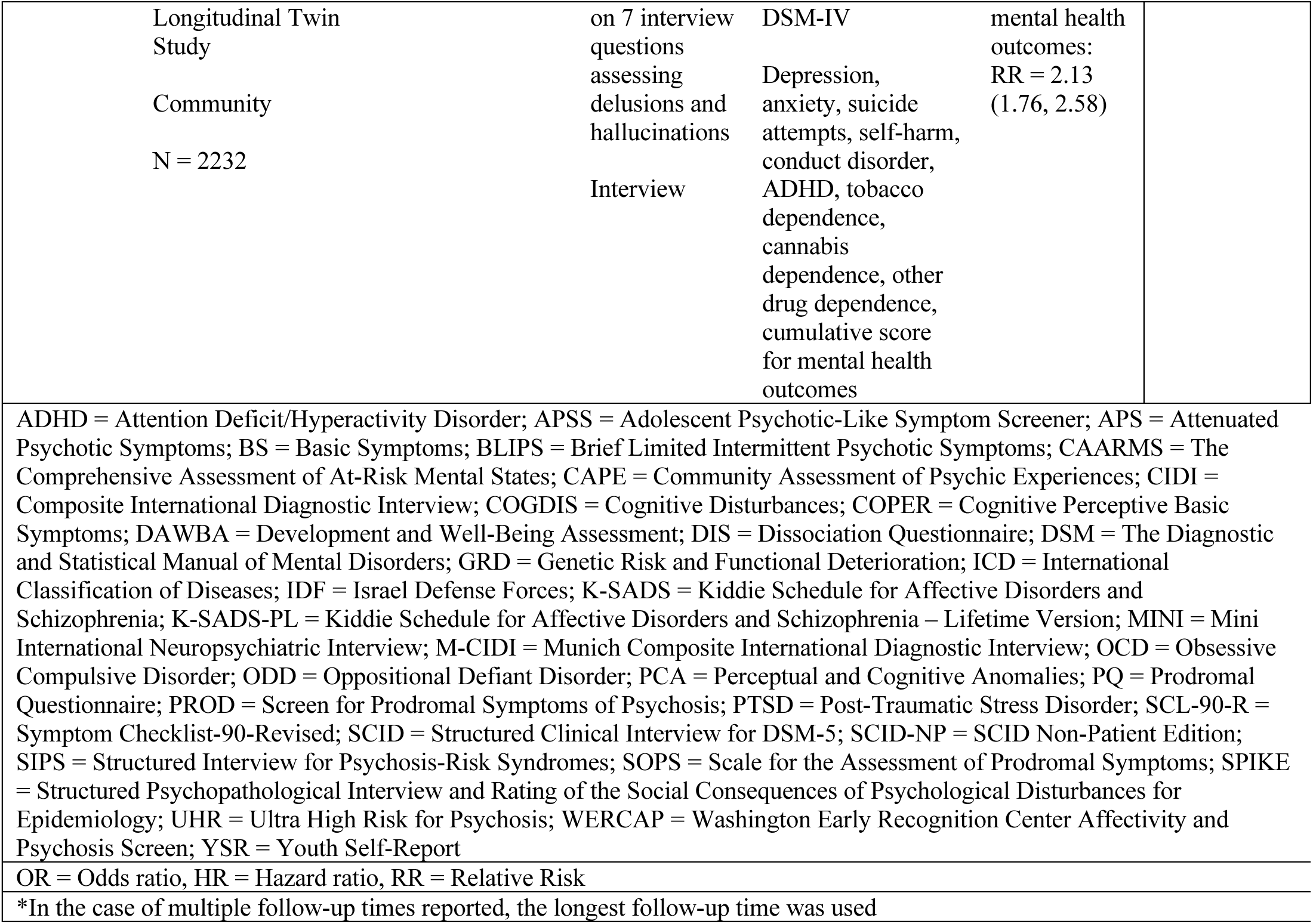
Descriptive summary of included studies.

**Table 2.** Pooled ORs and heterogeneity measures of the association between PSS and different sub categories of mental health disorder.

| Mental Health Disorder | Number of Studies | I<sup>2</sup> | Pooled OR (95% CI) |
| --- | --- | --- | --- |
| Psychotic | 10 | 60.93 | 3.11 (2.11 - 4.58) |
| Anxiety/Stress | 6 | 90.93 | 1.03 (0.55 - 1.83) |
| Substance Use | 8 | 79.44 | 1.88 (1.28 - 2.77) |
| Depressive | 8 | 93.76 | 1.32 (0.72 - 2.40) |

### Study Characteristics

Our search yielded 41 studies with an average age at baseline of 14.6 (SD = 4.0) and average follow up of 9.3 years (SD = 7.7, range=1-27 years). There were 12 studies that examined higher risk samples, including youth with a family history of schizophrenia, schizoaffective disorders, or bipolar disorders (k=6)^30,43,52,57,60,63^, youth meeting clinical high risk criteria (k=2)^61,62,64^, children/youth seeking mental health services in the absence of current or previous psychotic disorder diagnoses at baseline assessment (k=2)^45,55^, and psychiatric inpatients receiving mental health care for non-psychotic illnesses (k=1)^44^. Where more than one study used the same sample, we chose to include the study that had the most complete data for our analyses and largest sample size (Supplemental Figure 1). The remaining 29 studies come from 20 different community samples (Supplemental Text). A variety of PSS assessments were used. Eighteen studies used an interviewer-administered or rated questionnaire, twenty used validated self-report questionnaires, four studies developed their own questions to assess PSS presence/absence^35,53,57,70^, and three studies used both interview-based and self-report measures^32,52,66^.

Mental health diagnosis was assessed at follow up timepoints using the SCID^30,32,34,42,45,52,56,58,61,64^, DIS^31,33,48,50,53,66,70^, DISC^40^, National patient register^35,37,44,49,65,67^, CIDI^38,39,47,59^, DAWBA^41,68^, K-SADs^43,64^, MINI^46,62,69^, SPIKE^51^, SIPS and symptoms verified to meet criteria defined by DSM-IV and ICD-10 ^54,66^, and Present State Examination^63^.

## Meta-Analysis

### Association between PSS and subsequent mental health disorder diagnosis (OR Model 1)

The meta-analysis included 25 studies, of which, 16 examined mental health disorder outcomes and were used to investigate the relationship between an earlier measurement of childhood/adolescent PSS in the absence of diagnosable psychotic disorder diagnosis and later diagnosed mental health disorder (follow up ranging from 1-27 years after initial PSS assessment). Ten of these studies examined for any mental health disorder as the outcome^30,32,34,37,38,40–43,51^, while six examined for psychotic disorders, specifically, as the outcome of interest^31,44–46,53,59^. Six studies were excluded due to repeats in the study sample (described above). Two studies were identified as impactful outliers using studentized residuals and the leave-one-out analysis^52,54^ (Supplemental Figure 2). The results show that childhood/youth PSS were associated with a two-fold increased risk for diagnosis of any subsequent mental health disorder (OR = 2.07, CI = 1.61 - 2.66), with substantial heterogeneity among the studies analyzed (I^2^ = 86.92%, p < 0.0001). Visual inspection of the funnel plots (Supplemental Figure 3) and Egger’s regression test suggested asymmetry within the included studies on mental health disorders as outcomes (z = 3.72, p = 0.0002). To account for the observed asymmetry, we used a trim and fill method that yielded a slightly attenuated, but still statistically significant association between earlier documented PSS and later presence of any mental health disorder diagnosis (OR = 1.82, 95% CI 1.42–2.32). We conducted an additional sensitivity analysis in which we excluded any studies ^34,37,38,43^ that examined lifetime mental health disorder diagnosis to capture emergence of incident mental health disorder associated with baseline PSS and the results remained the same (OR = 2.09, 95% CI 1.51-2.89). The PAF was calculated from eleven studies, showing that childhood and adolescent PSS accounted for 38.98% of mental health disorders.

### Associations between PSS and subsequent psychotic disorder, anxiety and stress disorder, substance use disorder, and depressive disorder diagnosis

When our outcomes were narrowed to subcategories of mental health disorder, we determined there was a sufficient number of studies for a meta-analysis of the outcome of incident psychotic disorder (OR Model 2). There were 12 studies ^31,32,39,42,44–46,48,49,52–54^ that reported psychotic disorder diagnosis at follow-up, of which two studies were identified as impactful outliers^42,52^ (Supplemental Figure 4) and were removed from the analysis. Early documented PSS was associated with a three-fold increased risk for a future psychotic disorder diagnosis (OR = 3.11, CI = 2.11 - 4.58) with substantial between-study heterogeneity present (I^2^ = 60.93%, p < 0.0090). No statistically significant evidence of funnel plot asymmetry (Supplemental Figure 5) was observed, as confirmed by Egger’s regression test (z = 0.81, p = 0.42). The PAF was calculated from available data in ten studies and indicated that 40.69% of psychotic disorders can be attributed to childhood and adolescent PSS.

For other disorder outcomes with a sufficient number of studies to carry out meta-analyses, we observed an association between PSS and subsequent substance use disorder diagnosis (k=8, OR = 1.88, CI = 1.28 - 2.77). There was no significant association found between earlier recorded PSS with later depressive (k=8) or for pooled analysis of anxiety or stress disorders as outcomes (nonspecified anxiety disorders:k = 4, PTSD:k = 1, social phobia: k=1).

### Meta-regression

#### Analysis of Mental Health Disorder Diagnosis as Outcome (OR Model 1)

Given the substantial heterogeneity observed across studies, we investigated whether study-level characteristics moderated the association between PSS and subsequent mental health disorder diagnosis. We had sufficient data to examine the following moderators: PSS assessment type (self-report vs. interview-based questionnaires), sample type (clinical vs community), outcome type (any mental health disorders vs. psychotic disorders), sample size, average age at baseline, and follow-up time. None of these variables had a significant effect on the relationship between PSS and any mental health disorder outcome or helped to explain the heterogeneity observed in the literature (Supplemental Table 2). In order to further probe meta-regression findings, we conducted subgroup analyses examine whether associations between PSS and later mental health diagnosis varied for specific subgroups (community vs. higher-risk samples, average age<14 vs. average age>14, PSS assessed with an interview vs. self-report, follow-up time under 10 years vs. above 10 years). Pooled ORs, reported above, remained consistent within different subsamples (Supplemental Table 2).

#### Analysis of Psychotic Disorder Diagnosis as Outcome (OR Model 2)

In a mixed-effects meta-regression analysis of the 10 studies that reported psychotic disorders as an outcome, the included moderators significantly explained between-study variability in effect sizes. The differences across studies were largely accounted for by study quality and sample size, such that the identified OR between PSS and psychotic disorder in the current study became more stable when studies were rated as higher quality or had larger sample size (meta-analysis heterogeneity without moderators I^2^ = 60.93% versus with moderators I^2^ = 17.77%)(Supplemental Table 2).

## Discussion

This systematic review and meta-analysis extends prior findings of the mental health burden associated with PSS. Our findings, in the largest meta-analysis on the topic to date, confirm prior reports of a 2-fold increase in risk for an emergent or persistent mental health disorder diagnosis and a 3-fold increase in risk for incident psychotic disorder among children and youth who endorsed PSS at a previous timepoint. Most included studies that contributed to this finding used a brief self-report measure. Relative to previous meta-analyses, we were able to include more studies with a larger overall samples (k=16 vs prior largest k=7, n=27,100 vs prior largest n=1,571), a wider age range (5-23 years), a longer follow up duration (1-27 years), and more diverse sampling (e.g., clinical and community samples). Importantly, by including both community-based and at-risk or help seeking samples who may have baseline psychiatric diagnosis, we demonstrated for the first time that the links between PSS and subsequent mental health outcomes persist among different subsamples, including across varying levels of baseline clinical risk, and across reliance on interview versus self-reported PSS measurement.

While the increased risk observed in this meta-analysis is consistent with the literature, we observed lower effect sizes compared to previous meta-analyses. Prior work reported a threefold increased risk for any mental health disorder diagnosis^13^ and almost fourfold increased risk for psychotic disorders^13,14^. A key distinction between the present study and earlier reviews is that we included substantially more studies for both our meta-analysis of mental health disorders (k=16 vs prior k=7) and psychotic disorders (k=10 vs prior k=5) longitudinally, which may have resulted in more conservative and stable estimates. Meta-analyses integrating less data may be more susceptible to inflated effect sizes due to limitations of including individual studies^72^. Additionally, prior meta-analyses of this literature pooled all mental health disorder outcomes together^13^, whereas we made an effort to examine the association between prior PSS and specific subcategories of mental health diagnosis, where possible. Thus, prior observed odds ratios indicating a higher odd of subsequent mental health diagnosis among youth with PSS may reflect which disorders were included as outcomes (e.g., psychotic disorder or substance use disorders). These results support the growing suggestion from the literature that PSS in children and youth should not be considered developmentally normative phenomena but represent an important marker of psychiatric vulnerability, even when reported at just one timepoint. Our findings of specific risk for subsequent psychotic disorders among youth endorsing PSS emphasize that while PSS may be a relevant risk for mental health diagnosis transdiagnostically, targeted follow-up of identified PSS with assessment for CHR may be useful to identify individuals requiring specific monitoring and intervention to reduce the risk of developing a psychotic disorder. Given the low prevalence of individuals with first-episode psychosis that have prior contact with CHR services, PSS may be useful for identifying individuals who may otherwise go undetected ^73^. This perspective aligns with the clinical staging framework, which conceptualizes psychotic disorders as evolving along a continuum of severity and specificity over time^74^. Within this model, stages 0 or 1 identify individuals with family history or prodromal symptoms meeting CHR designation respectively. More widespread measurement of PSS may therefore help to identify and direct resources towards individuals at earlier stages of illness, in an effort to reduce risk of transition to psychotic disorder (i.e., stage 2+ of illness)^75,76^.

In our meta-regression analysis, our results suggested that the large heterogeneity found across studies examining the association between earlier reported PSS and subsequent mental health diagnosis was not explained by moderators that could be submitted to meta-regression based on available studies (i.e., clinical/community sample ascertainment, age of baseline assessment, length of study follow up time). In contrast, the heterogeneity that was found between studies that examined associations between earlier reported PSS and development of threshold symptoms for a later psychotic disorder was influenced by sample size and study quality. A substantial reduction in between study-heterogeneity was found when these variables were accounted for (I^2^ ∼18% vs. ∼60%), whereby greater sample size and better study quality was associated with a higher OR and more stable effect size. It is notable that the substantial reduction of between-study heterogeneity for the association between PSS and subsequent psychotic disorder diagnosis, found only when accounting for sample size and study quality, indirectly suggests that other potential moderators (e.g., age range of sample, community vs. clinical sample) do not have a substantial impact on between-study heterogeneity in the current literature. Thus, future work in this field would benefit from inclusion of larger samples, and improved study quality to increase confidence in PSS as a risk factor for subsequent emergence of a diagnosable psychotic disorder.

While our overall results indicated an association between PSS and later risk of subsequent mental health diagnosis, closer examination of the data available indicated that the extent to which PSS predicts the future development of non-psychotic disorders remains unclear. Specifically, our subcategory analyses indicated that PSS may provide more specific indication of prospective risk for developing threshold criteria for psychotic disorders and to a lesser extent substance use disorder, as opposed to depressive or anxiety/stress disorders. Previous studies have shown that individuals with PTSD symptoms are more likely to report PSS^77,78^. Specific attenuated psychotic experiences, such as delusions and hallucinations, have also been associated with an increased risk of a depressive disorder diagnosis^79^. Consistent with this, studies have also shown that youth with anxiety or depressive disorders are more likely to endorse PSS than individuals without mental health conditions^80^. Moreover, youth with PSS in the Philadelphia Neurodevelopmental Cohort (PNC) were more likely to also endorse symptoms of depression, mania, anxiety, and behavioural disturbances. Thus, while the presence of a prospective relationship between PSS and subsequent depressive or anxiety/stress diagnosis, as examined here, remains unclear, the prior literature has established that there is a concurrent risk to experience PSS in the context of a non-psychotic mental health diagnosis, such as depressive or anxiety/stress diagnoses. Our review also highlights the importance of further examination of whether there are important differences between PSS reported in community versus health seeking samples. Previous work has shown that youth aged 13-17 who access mental health services are at increased risk of being diagnosed with a psychotic or bipolar disorder at 28, and this risk is increased among youth requiring psychiatric hospital admissions^81^. These results underscore the clinical value of assessing psychosis risk in youth presenting with a range of mental health symptoms. The Toronto Adolescent and Youth (TAY) CAMH Cohort Study, focused on assessing PSS in a help-seeking sample of youth, will provide further insight into questions surrounding the importance of measuring PSS as a predictor of outcome in clinical youth samples^20^.

## Limitations

Several limitations should be noted. First, substantial heterogeneity was observed across studies included in the meta-analysis. This variability is likely attributable to differences in study design, quality, as well as PSS assessment used, follow-up duration, and baseline psychopathology assessments. In the meta-regression, PSS measures were broadly categorized as interview-based or self-report, which may have obscured important methodological distinctions. For example, a single self-report item assessing the presence of auditory hallucinations was grouped with a multi-item questionnaire with threshold-based scoring for meeting PSS criteria. This categorization followed Burton et. al., who reported a significant moderating effect of PSS assessment type within their meta-analysis that looked at psychotic disorder outcomes ^14^. They found that interview-based assessments were better at identifying risk for future psychotic disorder diagnosis, a finding which we did not detect in our analysis. Follow-up durations also varied, and many studies assessed lifetime prevalence at follow up, limiting the ability to disentangle the experience of PSS from timing of diagnosis onset. Finally, studies did not always systematically assess baseline general psychopathology nor confirm the absence of diagnosable psychiatric disorder (other than psychotic disorder) at the time of PSS assessment. Thus, the increased risk of meeting criteria for a subsequent mental health disorder diagnosis when earlier PSS is reported, as found here, could also represent increased risk of persistence of clinical symptoms meeting criteria for a later diagnosable disorder when earlier PSS is reported. Finally, the exclusion of studies that assessed psychotic symptoms using dimensional or non-diagnostic outcomes may have resulted in the omission of relevant findings. A number of studies measured transition to psychotic disorder, however few reported conversion rates to psychosis according to DSM/ICD criteria, especially studies conducted in community-based populations.

There are a number of additional factors that could have influenced our results, but for which insufficient data was available to evaluate them. For example, substance use has been associated with psychotic disorders, however we were not able to add this variable to our meta-regressions due to insufficient number of available studies with baseline substance use information. In addition, just three reviewed studies accounted for cannabis use when examining the association between PSS and future mental health disorder diagnosis. Among these, a healthy comparison group was available in two studies, with both suggesting that baseline PSS without cannabis use remained associated with an increased prospective risk of subsequent mental health disorder ^37^ or psychotic disorder^67^. Of note, in these studies, individuals with both PSS and cannabis use exhibited a twofold higher incidence of psychosis by the age of 30 compared to those with PSS alone^67^. Additionally, we were not able to examine whether repeated report or persistence of PSS over time may increase risk for later mental health diagnosis. Prior studies have shown that persistent PSS are associated with poorer mental health outcomes compared to transient or remitted symptoms^39,56^.

## Conclusion

This review demonstrates that a one-time endorsement of PSS in children and youth who do not meet threshold for a psychotic disorder at the time of assessment is a significant predictor of risk for subsequent mental health and specifically psychotic disorder diagnosis. This conclusion extends and enhances generalizability of prior results through careful inclusion of studies measuring prospective risk, expanding the age range to include the developmental period when peak prevalence of psychotic disorder occurs, and the integration of familial high-risk and help-seeking, as well as community samples. Thus, overall our results provide further confidence that incorporating a simple to use and easy to collect self-report measure of PSS can help to identify youth at increased risk for developing severe mental illness, across both clinical and non-clinical contexts.

## Data Availability

All data produced in the present study are available upon reasonable request to the authors

## Acknowledgments and Disclosures

KC currently receives funding from the CIHR, SSHRC, the Government of Ontario (Early Researcher Award), and the University of Toronto. DBC currently receives funding from the CIHR, the University of Toronto, the Cundill Centre for Child and Youth Depression, and the Margaret and Wallace McCain Centre for Child, Youth and Family Mental Health. GF currently receives funding from the CIHR, the CAMH Foundation, and the University of Toronto. NK currently receives funding from the PSI Foundation, CIHR, University of Toronto, Ontario Brain Institute, University of Toronto, CAMH AFP Innovation Fund, Brain Canada, the CAMH Foundation, and the Social Sciences and Humanities Research Council. ANV currently receives funding from the NIMH, the CIHR, the Canada Foundation for Innovation, the CAMH Foundation, the University of Toronto, Brain Canada, and Wellcome Trust. WW currently receives funding from the CIHR, the Canadian Centre on Substance Use and Addiction, the Ontario HIV Treatment Network, the Miner’s Lamp Innovation Fund, and the CAMH Foundation. SHA currently receives funding from the National Institute of Mental Health (NIMH), Canadian Institutes of Health Research (CIHR), the Canada Research Chairs program, and the CAMH Foundation. EWD currently receives funding from the NIMH, CIHR, the CAMH Foundation, Wellcome Trust, and Department of Psychiatry Academic Scholar’s Award. All other authors report no biomedical financial interests or potential conflicts of interest.

