## Supplement for "Systematic Review and Meta-Analysis: Do Youth-Reported Psychosis Symptoms Predict Later Mental Health Diagnosis?"

Study Selection Supplemental Text

Our search yielded 41 studies with an average age at baseline of 14.6 (SD = 4.0) and average follow up of 9.3 years (SD = 7.7, range=1-27 years). There were 11 studies that examined clinical samples, including youth with a family history of schizophrenia, schizoaffective disorders, or bipolar disorders (k=6)[30,43,52,57,60,63], high risk youth referred for assessments by healthcare professionals or concerned family members (k=2)[62,64], children/youth seeking mental health services in the absence of current or previous psychotic disorder diagnoses at baseline assessment (k=2)[45,55], and psychiatric inpatients receiving mental health care for non-psychotic illnesses (k=1)[44]. Where more than one study used the same sample, we chose to include the study that had the most complete data for our analyses and largest sample size (Supplemental Figure 1). The remaining 30 studies come from 20 different community samples (Supplemental Figure). Of these, nine used data from population-based birth cohorts[33,37,40,48–50,54,67,68], with two additional studies using twin data from birth cohorts[35,70]. Five studies recruited from primary schools[31,34,36,41,58], while three recruited adolescents from high school or secondary schools[46,66,69]. Two studies consisted of population-based samples of youth receiving medical care at the Children's Hospital of Philadelphia (CHOP)[32,56], and three obtained a random population of youth from the general population[38,39,59]. One study sampled college undergraduate students enrolled in a psychology course[42], two examined offspring of pregnant women receiving care in a public hospital in Australia[47,53], and one used a web-based questionnaire to identify community participants meeting a PSS symptom threshold[61]. One study recruited participants in Zurich using male conscription data and female electoral registrar data[51], and one study used data from mental health examinations conducted by the Israeli Defense Forces[65].

A variety of PSS assessments were used. Eighteen studies used an interviewer-administered or rated questionnaire(e.g., SIPS/SOPS[30,32,55,62–64,66], K-SADS[32,34,43,44,68], DISC[33,40,48,50], SPIKE[51], Psychosis-Like Symptom Interview[54], Brief Psychiatric Rating Scale (BPRS)[59], CAARMS[61], and IDF Mental Health Examination[65]). Twenty used self-report questionnaires (APSS[36,58], PROD[37,49,67], YSR[31,38,47], SCL-90-R[39], CAPE[41,46,69], Chapman scale[42,60], PQ [45], PRIME[56]). Four studies developed their own questions to assess PSS presence/absence[35,53,57,70]. Three studies used both interview-based and self-report measures[32,52,66].

Mental health diagnosis was assessed at follow up timepoints using the SCID[30,32,34,42,45,52,56,58,61,64], DIS[31,33,48,50,53,66,70], DISC[40], National patient register[35,37,44,49,65,67], CIDI[38,39,47,59], DAWBA[41,68], K-SADs[43,64], MINI[46,62,69], SPIKE[51], SIPS and symptoms verified to meet criteria defined by DSM-IV and ICD-10 [54,66] , and Present State Examination[63].

**Supplemental Figure 1. Overlapping Study Sample Selection**

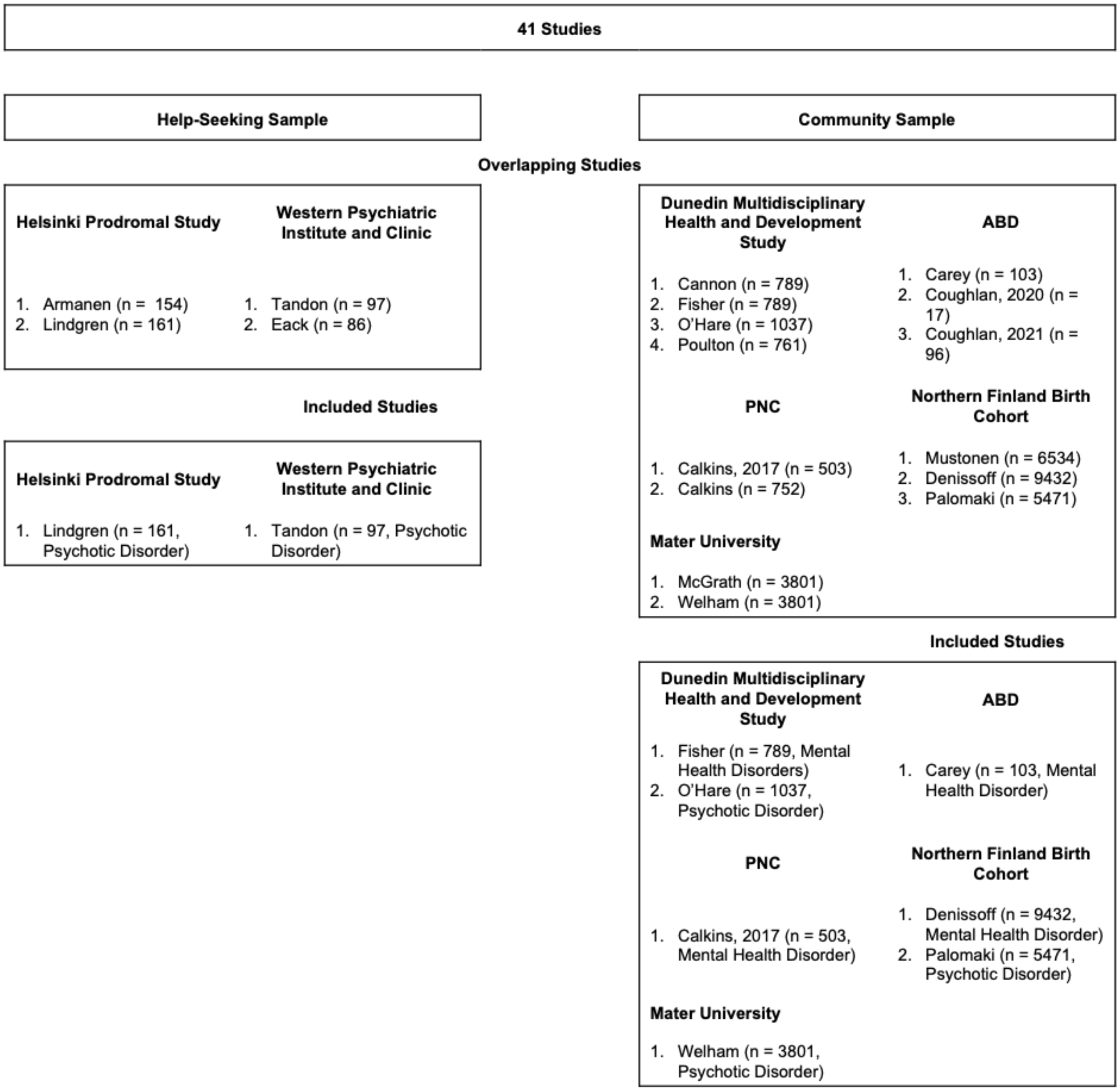


**Supplemental Figure 2. Forest Plot of Odds Ratios for Mental Health Disorders with Outliers**

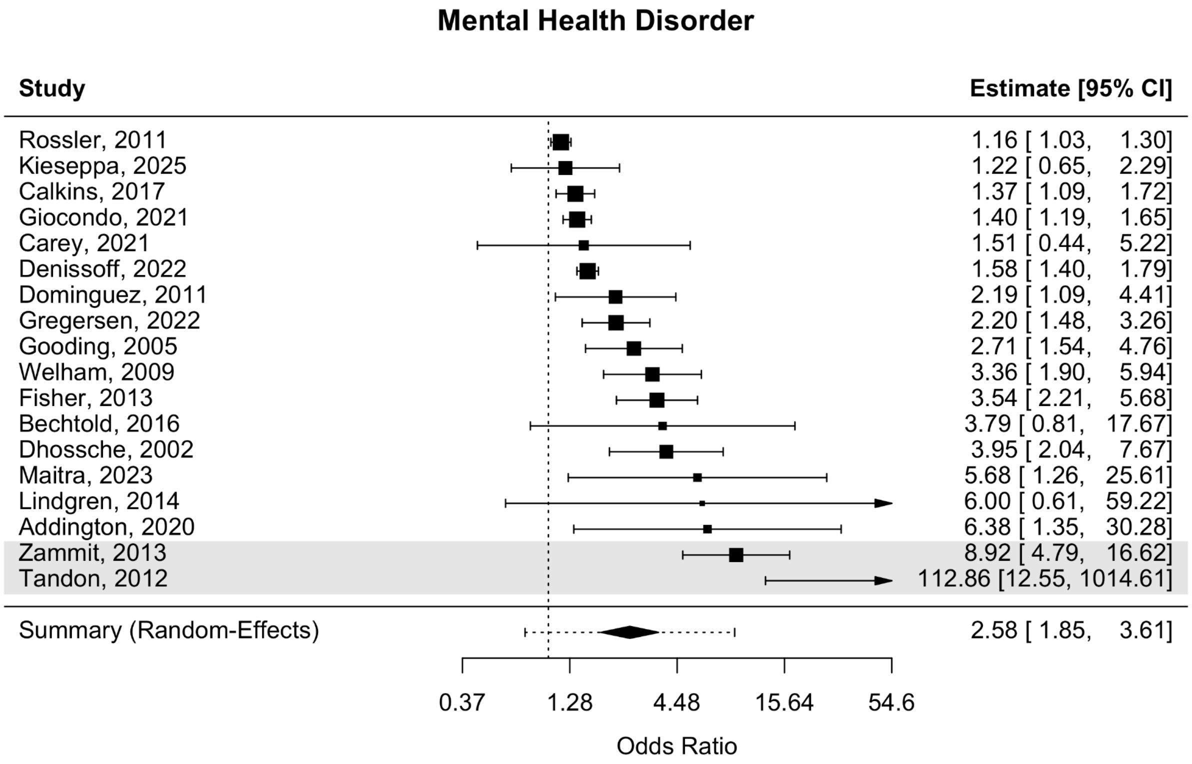


**Supplemental Figure 3. Funnel Plot of Mental Health Disorders as the Outcome**

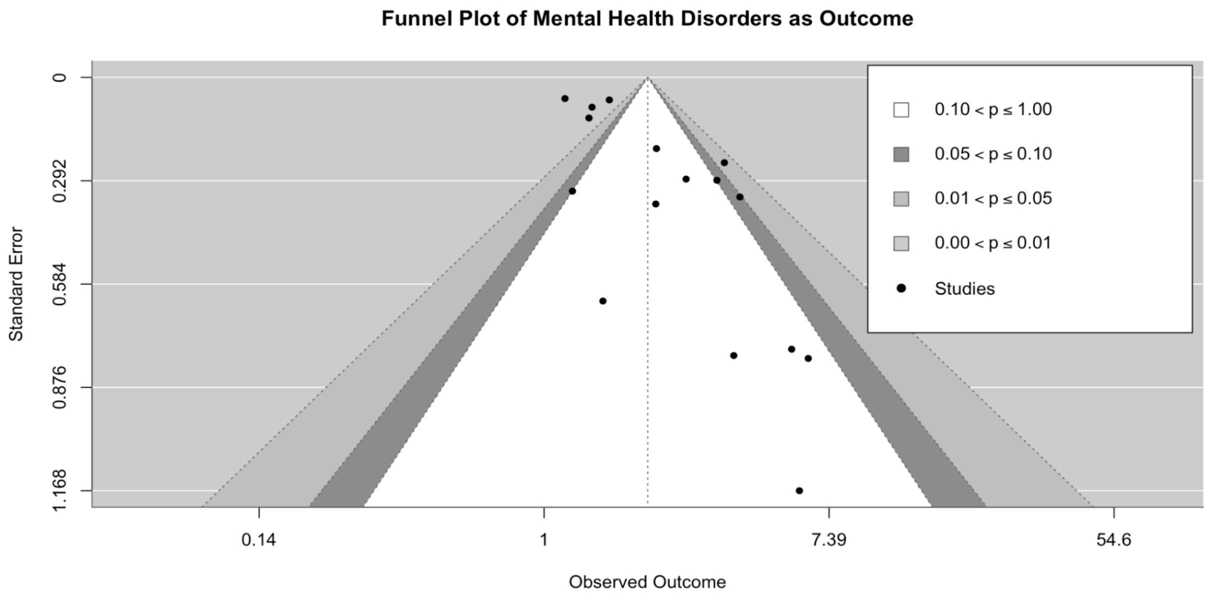


**Supplemental Figure 4. Forest Plot of Odds Ratios for Psychotic Disorders with Outliers**

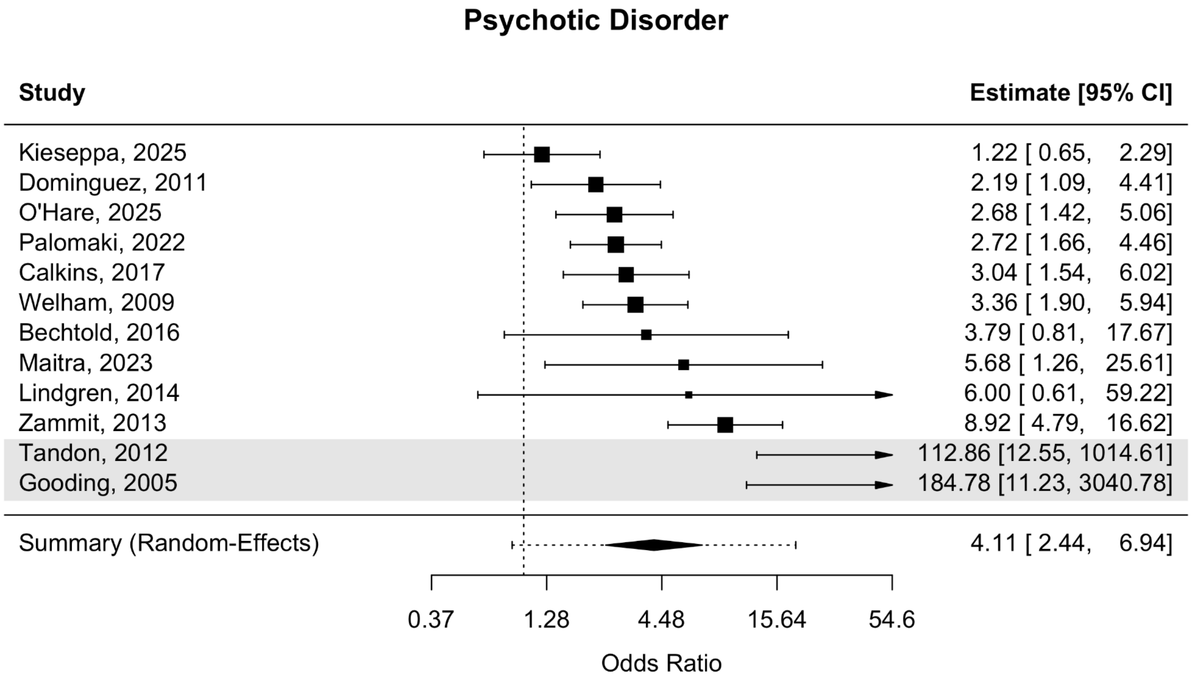


**Supplemental Figure 5. Funnel Plot of Psychotic Disorders as the Outcome**

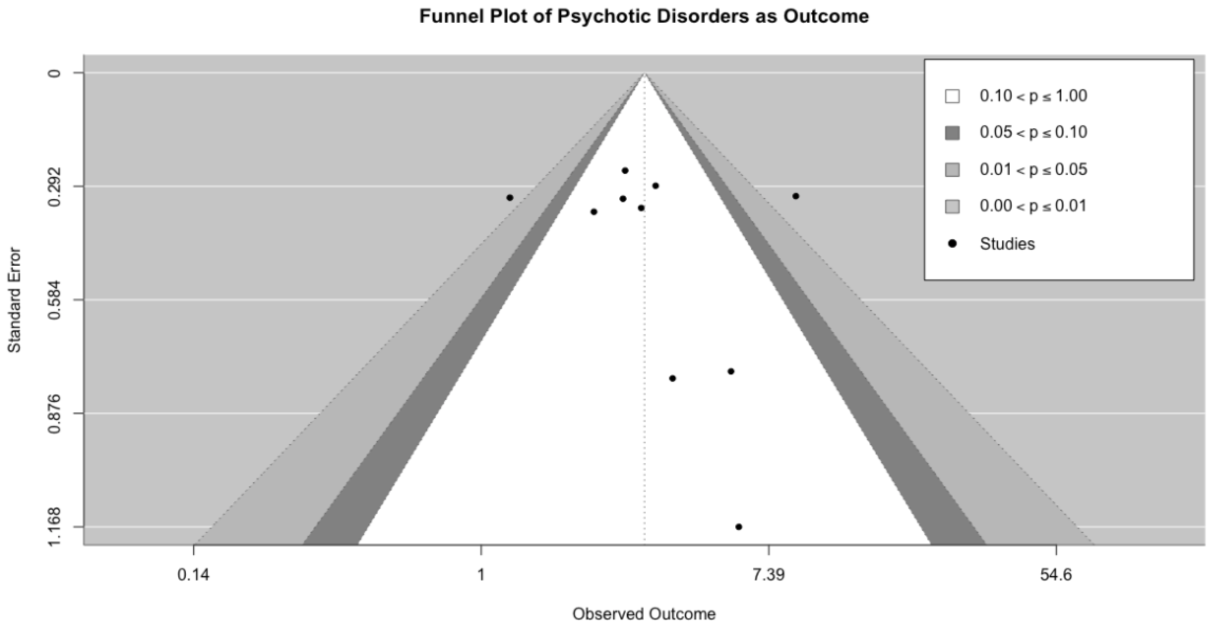


**Supplemental Figure 6. Bubble plot of Meta-Regression of Effect Size by Sample Size**

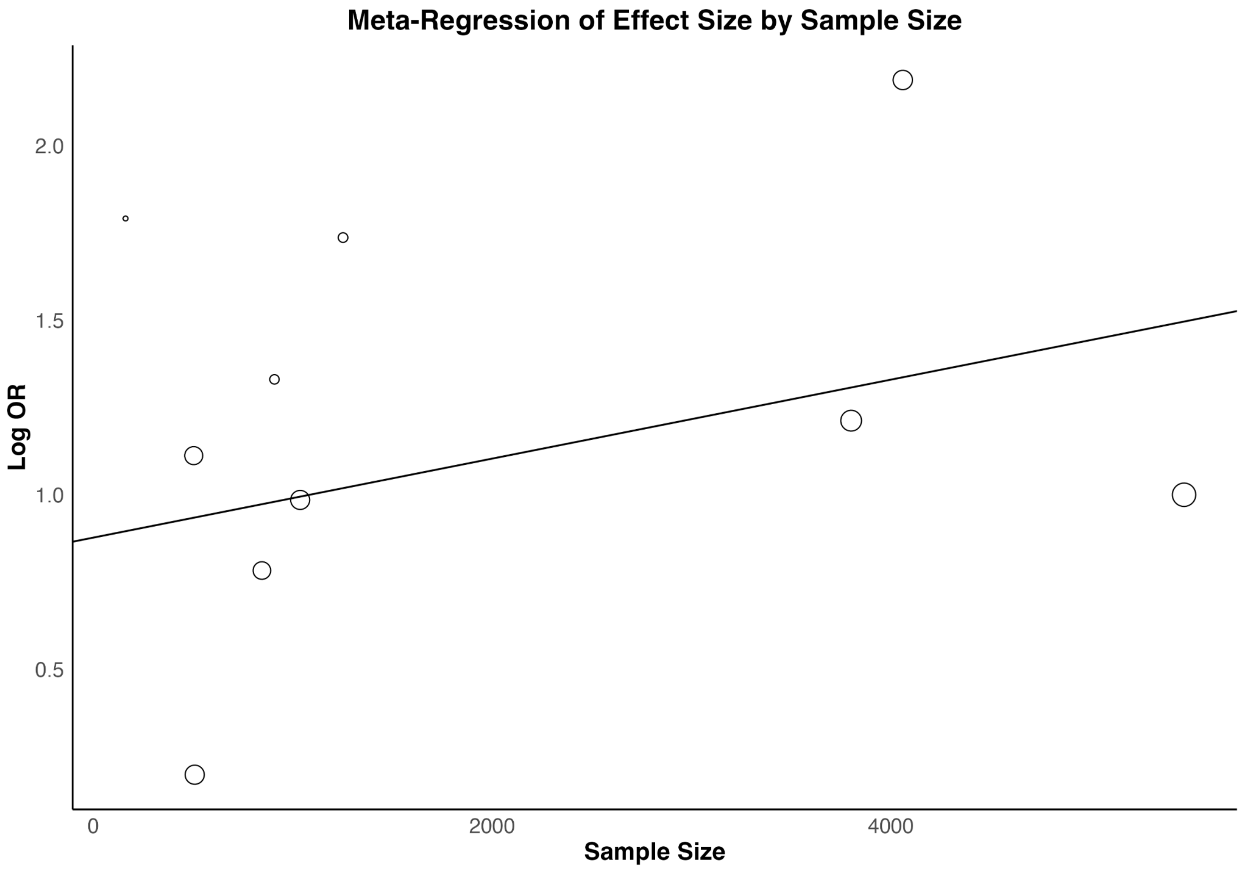


**Supplemental Figure 7. Bubble plot of Meta-Regression of Effect Size by Quality of Study**

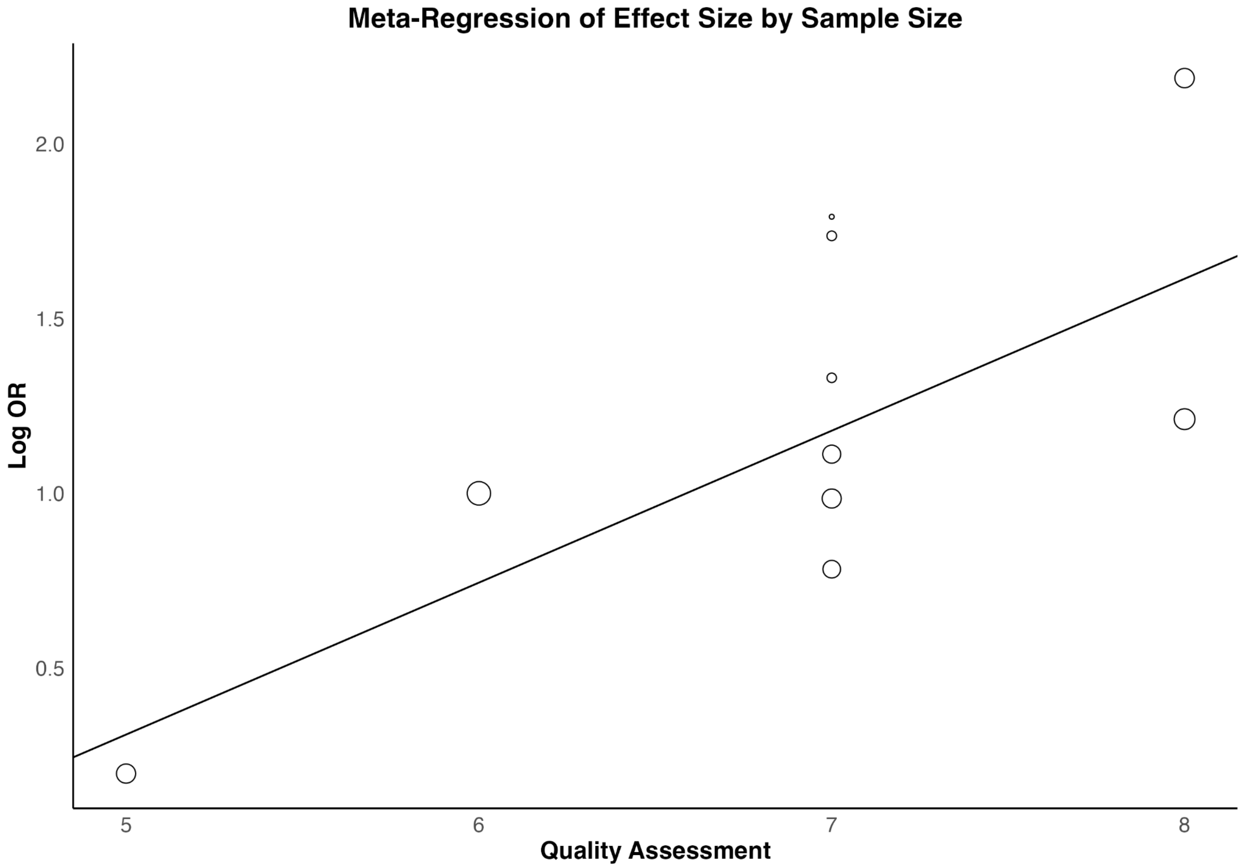


### Supplemental Table 1: Modified Newcastle-Ottawa Scale

| Score | Description |
|---|---|
| | ***Adequacy of the PSS definition*** |
| 1 | Validated assessment, dichotomous assessment, clinician rating |
| 0 | No description of PSS assessment |
| | ***Depth of sample characterization*** |
| 1 | Includes information such as family history, substance abuse, and comorbidities |
| 0 | No additional sample information provided |
| | ***Representativeness of the PSS cohort*** |
| 1 | Representative of the youth population |
| 0 | Select sample only (i.e., restricted to a specific sex/gender, race/ethnicity, high-risk cohorts, non-help seeking etc.) |
| | ***Availability of controls (Non-PSS group)*** |
| 1 | A control group is available |
| 0 | No |

| | |
|---|---|
| | ***Assessment of mental health disorder outcome*** |
| 1 | DSM/ICD based diagnosis for psychotic disorder or mental health disorder |
| 0 | No description of disorder outcome |
| | ***Other outcomes assessed at follow up*** |
| 1 | Did the study assess other mental health and functional outcomes |
| 0 | No other outcomes addressed |
| | ***Statistical test*** |
| 1 | The statistical test used to analyze the data is clearly described and appropriate, and the measurement of the association is presented, including confidence intervals and/or the probability level (p-value) |
| 0 | The statistical test is not appropriate, not described, or incomplete |
| | ***Adequacy of follow-up studies*** |
| 1 | Sufficient follow up data on controls and PSS with diagnosis for both at follow up |
| 0 | Incomplete follow up data or follow up not sufficiently reported |
| **8** | *7+/8 good, 5+/8 moderate, 4-/8 poor* |

**Supplemental Table 2: Sub-group analysis with mental health and psychotic disorder outcome**

| | Mental Health Disorder | | | Psychotic Disorder | | |
|---|---|---|---|---|---|---|
| **Sub-Group Analysis** | **Number of Studies** | **$I^2$** | **Pooled OR (95% CI)** | **Number of Studies** | **$I^2$** | **Pooled OR (95% CI)** |
| **Sample Type** | | | | | | |
| Community | 12 | 90.36 | 2.07(1.55, 2.76) | 8 | 42.14 | 1.82(0.47,7.05) |
| Higher-Risk | 4 | 50.14 | 2.16(1.17, 4.00) | 2 | 48.37 | 3.48(2.44,4.95) |
| **Average Age** | | | | | | |
| Under 14 | 6 | 72.70 | 2.31(1.56, 3.43) | 3 | 60.97 | 5.11(2.46,10.60) |
| Above 14 | 10 | 88.52 | 1.94107(1.39, 2.69) | 7 | 18.05 | 2.41(1.77,3.27) |
| **PSS Assessment Type** | | | | | | |

| | | | | | | |
|---|---|---|---|---|---|---|
| Interview | 7 | 85.60 | 1.78 (1.22,2.61) | 4 | 84.24 | 3.07(1.37,6.89) |
| Self-Report | 9 | 81.75 | 2.37 (1.68,3.34) | 6 | 0 | 2.95(2.16,4.03) |
| **Follow-up Time** | | | | | | |
| Under 10 | 9 | 74.29 | 2.17 (1.56,3.01) | 5 | 58.93 | 4.06(2.13,7.74) |
| Above 10 | 7 | 92.25 | 1.96(1.29,2.98) | 5 | 45.10 | 2.51(1.68,3.74) |
| Meta Regression Results: Age + Follow up time + Sample size + QA | | | | | | |
| | $I^2 = 77.89\%$ | | $R^2$=0.00% | $I^2 = 17.77\%$ | | $R^2$=83.41% |

**Supplemental Table 3. PRISMA Checklist**

| Section and Topic | Item # | Checklist item | Location where item is reported |
|---|---|---|---|
| **TITLE** | | | |
| Title | 1 | Identify the report as a systematic review. | Title page |
| **ABSTRACT** | | | |
| Abstract | 2 | See the PRISMA 2020 for Abstracts checklist. | Page 1 |
| **INTRODUCTION** | | | |
| Rationale | 3 | Describe the rationale for the review in the context of existing knowledge. | Page 1/2 |
| Objectives | 4 | Provide an explicit statement of the objective(s) or question(s) the review addresses. | Page 2 |
| **METHODS** | | | |
| Eligibility criteria | 5 | Specify the inclusion and exclusion criteria for the review and how studies were grouped for the syntheses. | Page 4 |
| Information sources | 6 | Specify all databases, registers, websites, organisations, reference lists and other sources searched or consulted to identify studies. Specify the date when each source was last searched or consulted. | Page 3 |
| Search strategy | 7 | Present the full search strategies for all databases, registers and websites, including any filters and limits used. | Page 3 |
| Selection process | 8 | Specify the methods used to decide whether a study met the inclusion criteria of the review, including how many reviewers screened each record and each report retrieved, whether they worked independently, and if applicable, details of automation tools used in the process. | Page 4 |

| | | | |
|---|---|---|---|
| Data collection process | 9 | Specify the methods used to collect data from reports, including how many reviewers collected data from each report, whether they worked independently, any processes for obtaining or confirming data from study investigators, and if applicable, details of automation tools used in the process. | Page 4 |
| Data items | 10 a | List and define all outcomes for which data were sought. Specify whether all results that were compatible with each outcome domain in each study were sought (e.g. for all measures, time points, analyses), and if not, the methods used to decide which results to collect. | Page 5 |
| | 10 b | List and define all other variables for which data were sought (e.g. participant and intervention characteristics, funding sources). Describe any assumptions made about any missing or unclear information. | Page 5 |
| Study risk of bias assessmen t | 11 | Specify the methods used to assess risk of bias in the included studies, including details of the tool(s) used, how many reviewers assessed each study and whether they worked independently, and if applicable, details of automation tools used in the process. | Page 4 |
| Effect measures | 12 | Specify for each outcome the effect measure(s) (e.g. risk ratio, mean difference) used in the synthesis or presentation of results. | Page 5 |
| Synthesis methods | 13 a | Describe the processes used to decide which studies were eligible for each synthesis (e.g. tabulating the study intervention characteristics and comparing against the planned groups for each synthesis (item #5)). | Page 4 |
| | 13 b | Describe any methods required to prepare the data for presentation or synthesis, such as handling of missing summary statistics, or data conversions. | Page 5 |
| | 13 c | Describe any methods used to tabulate or visually display results of individual studies and syntheses. | Page 5 |
| | 13 d | Describe any methods used to synthesize results and provide a rationale for the choice(s). If meta-analysis was performed, describe the model(s), method(s) to identify the presence and extent of statistical heterogeneity, and software package(s) used. | Page 6 |
| | 13 e | Describe any methods used to explore possible causes of heterogeneity among study results (e.g. subgroup analysis, meta-regression). | Page 6 |
| | 13f | Describe any sensitivity analyses conducted to assess robustness of the synthesized results. | Page 5 |
| Reporting bias assessmen t | 14 | Describe any methods used to assess risk of bias due to missing results in a synthesis (arising from reporting biases). | Page 4 |
| Certainty assessmen t | 15 | Describe any methods used to assess certainty (or confidence) in the body of evidence for an outcome. | Page 5 |
| **RESULTS** | | | |
| Study selection | 16 a | Describe the results of the search and selection process, from the number of records identified in the search to the number of studies included in the review, ideally using a flow diagram. | Page 6 |

| | | | |
|---|---|---|---|
| | 16 b | Cite studies that might appear to meet the inclusion criteria, but which were excluded, and explain why they were excluded. | Page 6 |
| Study characteristics | 17 | Cite each included study and present its characteristics. | Table 1 |
| Risk of bias in studies | 18 | Present assessments of risk of bias for each included study. | Table 1 |
| Results of individual studies | 19 | For all outcomes, present, for each study: (a) summary statistics for each group (where appropriate) and (b) an effect estimate and its precision (e.g. confidence/credible interval), ideally using structured tables or plots. | Table 1 and Figure 2 and 3 |
| Results of syntheses | 20 a | For each synthesis, briefly summarise the characteristics and risk of bias among contributing studies. | Table 1 |
| | 20 b | Present results of all statistical syntheses conducted. If meta-analysis was done, present for each the summary estimate and its precision (e.g. confidence/credible interval) and measures of statistical heterogeneity. If comparing groups, describe the direction of the effect. | Figure 2 and Figure 3 |
| | 20 c | Present results of all investigations of possible causes of heterogeneity among study results. | Page 8 |
| | 20 d | Present results of all sensitivity analyses conducted to assess the robustness of the synthesized results. | Supplemental figures 2 and 4 |
| Reporting biases | 21 | Present assessments of risk of bias due to missing results (arising from reporting biases) for each synthesis assessed. | Table 1 |
| Certainty of evidence | 22 | Present assessments of certainty (or confidence) in the body of evidence for each outcome assessed. | Figures 2 and 3 |
| **DISCUSSION** | | | |
| Discussion | 23 a | Provide a general interpretation of the results in the context of other evidence. | Page 8-11 |
| | 23 b | Discuss any limitations of the evidence included in the review. | Page 11 |
| | 23 c | Discuss any limitations of the review processes used. | Page 11 |
| | 23 d | Discuss implications of the results for practice, policy, and future research. | Page 12 |
| **OTHER INFORMATION** | | | |
| | 24 a | Provide registration information for the review, including register name and registration number, or state that the review was not registered. | Page 3 |

| | | | |
|---|---|---|---|
| Registratio n and protocol | 24 b | Indicate where the review protocol can be accessed, or state that a protocol was not prepared. | Page 3 |
| | 24 c | Describe and explain any amendments to information provided at registration or in the protocol. | Page 3 |
| Support | 25 | Describe sources of financial or non-financial support for the review, and the role of the funders or sponsors in the review. | Title Page |
| Competing interests | 26 | Declare any competing interests of review authors. | Title Page |
| Availability of data, code and other materials | 27 | Report which of the following are publicly available and where they can be found: template data collection forms; data extracted from included studies; data used for all analyses; analytic code; any other materials used in the review. | Attache d to submis sion |
